# Parental education and income are linked to offspring cortical brain structure and psychopathology at 9-11 years

**DOI:** 10.1101/2023.02.16.23286027

**Authors:** Linn B. Norbom, Jaroslav Rokicki, Espen M. Eilertsen, Thea Wiker, Jamie Hanson, Andreas Dahl, Dag Alnæs, Sara Fernández-Cabello, Dani Beck, Ingrid Agartz, Ole A. Andreassen, Lars T. Westlye, Christian K. Tamnes

**Affiliations:** PROMENTA Research Center, Department of Psychology, University of Oslo, Norway; NORMENT, Institute of Clinical Medicine, University of Oslo, Norway; Department of Psychiatric Research, Diakonhjemmet Hospital, Oslo, Norway; Centre of Research and Education in Forensic Psychiatry, Oslo University Hospital, Oslo, Norway; Learning Research and Development Center University of Pittsburgh, USA; Department of Psychology, University of Pittsburgh, USA; Department of Psychology, University of Oslo, Norway; Department of Psychology, Pedagogy and Law, Kristiania University College, Norway; K.G Jebsen Center for Neurodevelopmental Disorders, University of Oslo, Norway; NORMENT, Division of Mental Health and Addiction, Oslo University Hospital & Institute of Clinical Medicine, University of Oslo, Norway; Centre for Psychiatry Research, Department of Clinical Neuroscience, Karolinska Institutet & Stockholm Health Care Services, Stockholm Region, Stockholm, Sweden

## Abstract

**Background:** A child’s socioeconomic environment can shape central aspects of their life, including vulnerability to mental disorders. Negative environmental influences in youth may interfere with the extensive and dynamic brain development occurring at this time. Indeed, there are numerous yet diverging reports of associations between parental socioeconomic status (SES) and child cortical brain morphometry. Most of these studies have used single metric- or unimodal analyses of standard cortical morphometry that downplay the probable scenario where numerous biological pathways *in sum* account for SES-related cortical differences in youth.

**Methods:** To comprehensively capture such variability, using data from 9758 children aged 8.9-11.1 years from the ABCD Study^®^, we employed linked independent component analysis (LICA) and fused vertex-wise cortical thickness, surface area, curvature and grey-/white-matter contrast (GWC). LICA revealed 70 uni- and multimodal components. We then assessed the linear relationships between parental education, parental income and each of the cortical components, controlling for age, sex, genetic ancestry, and family relatedness. We also assessed whether cortical structure moderated the negative relationships between parental SES and child general psychopathology.

**Results:** Parental education and income were both associated with larger surface area and higher GWC globally, in addition to local increases in surface area and to a lesser extent bidirectional GWC and cortical thickness patterns. The negative relation between parental income and child psychopathology were attenuated in children with a multimodal pattern of larger frontal- and smaller occipital surface area, and lower medial occipital thickness and GWC.

**Conclusion:** Structural brain MRI is sensitive to SES diversity in childhood, with GWC emerging as a particularly relevant marker together with surface area. In low-income families, having a more developed cortex across MRI metrics, appears beneficial for mental health.

## Introduction

The socioeconomic environment of a child can shape many central aspects of their life, including life expectancy, present and prospective cognitive abilities, school performance, and susceptibility for mental health struggles (Thomas & Coecke, 2023). Childhood is also marked by extensive brain development. These neuronal optimization processes, caused by genetic and environmental factors in complex interplay, are indirectly detectable by magnetic resonance imaging (MRI) (Jernigan et al., 2016; Lebel & Deoni, 2018; Norbom et al., 2021). Although brain plasticity fosters adaptation and learning, by the same token, environmental variables including socioeconomic status (SES) can affect brain development and influence the risk of mental disorders (Dearing et al., 2006; Farah, 2017; Letourneau et al., 2013).

As theorized by sociologist Pierre Bourdieu, SES is a complex dimensional construct used to assess social (connections), cultural (skills, knowledge and education), symbolic (prestige), and financial capital (Bourdieu, 2011).Within neuroscience, SES is often assessed by material gains like income, and non-material gains such as education and occupation (Long & Renbarger, 2023). This tradition has been critiqued and expanded to include subjective and individual experiences of social class (W. M. Liu et al., 2004), and cultural knowledge and abilities of marginalized minority groups that often go unrecognized (Yosso, 2005).

Low parental SES is associated with a broad array of negative outcomes in children. This includes lower cognitive abilities, a discrepancy that widens across childhood (Duyme et al., 1999; von Stumm & Plomin, 2015; Zhang et al., 2020), and poorer academic achievements (Sirin, 2005). Youths from lower SES families are also 2–3 times more likely to suffer from mental health problems than their higher SES peers (Letourneau et al., 2013; Reiss, 2013). Although the association between parental SES and youth psychopathology is established, how the brain affects these relations, remain poorly understood. SES is also heritable (Hill et al., 2019; Ørstavik et al., 2014; Tambs et al., 2012) complicating the causal relations with brain and mental health.

Childhood is a central period for brain maturation, involving multiple biological processes that show spatial and temporal heterogeneity across tissue types, metrics, and individuals (Jernigan et al., 2016; Lebel & Deoni, 2018; Norbom et al., 2021). The cerebral cortex shows a particularly lengthy developmental trajectory, with protracted decreases in apparent thickness and early increases in surface area and curvature (Norbom et al., 2021; Sydnor et al., 2021). Beyond morphometry, a prominent feature of youth development is an increase in cortical brightness. Variations in cortical brightness can be assessed through T1-weighted (T1w) intensity metrics such as the grey/white-matter contrast (GWC) (Salat et al., 2009). As cholesterol in myelin is a major determinant of the T1w-signal (Koenig, 1991; Koenig et al., 1990), GWC has been suggested as a viable proxy for intracortical myelination (Jørgensen et al., 2016), a crucial feature of postnatal brain development, allowing for efficient signal transmission and structural support (Bartzokis, 2012; Baumann & Pham-Dinh, 2001; S. Liu et al., 2019; Waxman & Bennett, 1972).

There are several reports of associations between parental SES and child cortical structure (Khundrakpam et al., 2020; Noble et al., 2015; Piccolo et al., 2016; Rakesh et al., 2022; Tomasi & Volkow, 2021). Findings are discrepant both in terms of the sensitivity of different imaging modalities, and the direction. Still, a recent comprehensive review of 71 studies pointed to quite consistent positive relations between parental SES and global as well as frontal surface area in childhood (Rakesh & Whittle, 2021). The review found no studies employing intensity metrics like GWC, or cortical curvature, but a single study on the related metric gyrification. The general divergence in findings could partly be explained by varying SES measures and distinct SES subfactor - cortical metric relationships (Farah, 2017; Rakesh & Whittle, 2021). Also, due to convention and availability, many recent studies use imaging ROI’s from Desikan-Killiany or the Destrieux atlas (Desikan et al., 2006; Destrieux et al., 2010). These atlases are not based on cyto- or myeloarchitecture, which are the main proposed neurobiological drivers of cortical thickness, area and GWC, but instead on gyral folding. Rather than adhering to a priori divisions, one could perform a data driven reduction of the vertex-wise data. A related challenge is that most studies have assessed a single-, or a few selected morphometric measures separately. This downplays the probable scenario of complex constructs like SES affecting numerous biological pathways with unique genetic and environmental determinants (Hogstrom et al., 2013; Rakic, 1988; Strike et al., 2019) that in sum underlie the SES – cortical structure relationship.

Multivariate imaging approaches (Groves et al., 2011; Miller et al., 2016) can co-model several sources of variability, which could increase effect sizes and improve neurobiological interpretation. For instance, while an independent component analysis (ICA) decomposes a signal into its constituent parts, linked ICA (LICA) can simultaneously “link”, or model common features across modalities irrespective of the units’ signal- and contrast-to-noise ratios and spatial smoothness (Groves et al., 2011). LICA studies have reported unique structural patterns sensitive to brain development and psychopathology (Groves et al., 2012; Norbom et al., 2020; Wolfers et al., 2017). Moreover, a recent study using canonical correlation analysis (CCA) reported common modes capturing SES factors and patterns of cortical morphometry, including sulcal depth (Alnæs et al., 2020). Thus, while it is reasonable to use multimodal reduction approaches that go beyond standard morphometry by including cortical curvature and GWC for a comprehensive assessment of the parental SES and child cortical structure relationship, this has not previously been done.

Using data obtained from 9758 children aged 8.9-11.1 years from the Adolescent Brain Cognitive Development (ABCD) Study^®^, we used LICA to perform multimodal fusion of vertex-wise cortical thickness, surface area, curvature and GWC. We then assessed the linear relation between parental education and parental income and each LICA component, and whether components could moderate the negative relationship between parental SES and general psychopathology. We hypothesized that parental education and income would show positive associations with global and frontal surface area (Noble et al., 2015; Rakesh & Whittle, 2021; Thomas & Coecke, 2023), and with widespread GWC for parental education (Norbom et al., 2022). We expected no parental SES - cortical thickness relations (Norbom et al., 2022; Rakesh & Whittle, 2021), and had no specific hypotheses pertaining to curvature or multimodal coupling.

## Materials and Methods

### Participants

Data was acquired from the ABCD Study^®^ using the curated annual release 4.0 (https://data-archive.nimh.nih.gov/abcd), with further detail described in the Supplemental Information (SI). The ABCD study^®^ consists of data from almost 12,000 children, aged approximately 9-10 years at study inclusion, as well as their parents. The data is obtained across 21 sites in the United States of America and includes detailed demographic, genetic, behavioral and neuroimaging data that will be collected for a decade (Feldstein Ewing et al., 2018).

The current study was based on the baseline assessment of the ABCD study^®^. From 11,876 participants, 8 children had partly, or completely missing Child Behavior Checklist (CBCL) raw scores and were therefore excluded. 14 children with missing educational data for both parents and 631 children with completely missing parental income information were also excluded. Another 732 children were excluded due to missing genetic ancestry data. On the imaging side we had FreeSurfer processed data for 11591 individuals. Of these 535 individuals did not pass MRI quality control or had missing data (see below) and were therefore excluded. 565 subjects were excluded during the intersection of individuals with complete demographics and complete neuroimaging resulting in a final sample size of 9758 participants (5124 females) aged 8.9-11.1 years (mean =9.9, SD =0.6), including 18 triplets and 1662 mono- and dizygotic twins.

### Measurement of socioeconomic status

Socioeconomic information was reported by a parent or guardian on behalf of themselves and a partner if relevant, by completing the “ABCD parental demographics survey”.

Parental education was assessed with the question “What is the highest grade/level/degree you have completed or received” ranging from 0 = Never attended/Kindergarten only, to 21 = Doctoral degree. We recoded this variable to years of total education as described in SI and defined parental education as the highest educational score of either the reporting parent or their partner.

Parental income was assessed with the question “How much did you earn, before taxes and other deductions, during the past 12 months?”, ranging from 1 = Less than $5.000, to 10 = $200.000 or greater. Total family income was assessed with the question “What is your total combined family income for the past 12 months?” using identical scoring. We recoded these variables using the median of each bracket as described in the SI and defined parental income as the highest number available from parent, partner, or combined income. The distributions of raw SES scores are shown in SI Figure 1.

### Assessment of general psychopathology

Child psychopathology was assessed using the CBCL (Achenbach & Ruffle, 2000), which is a widely used caregiver report for identifying behavioral and emotional problems in children. The CBCL contains 119 items pertaining to particular behaviors, and the reporter must assess, on a 3-point scale, to which extent these are characteristic of the child during the past six months. We calculated an overarching “p-factor” in accordance with “the general factor of psychopathology model 7” from Clark et al. (2021) where items were first grouped into three lower order factors, namely internalizing, externalizing and attention problems before being summed to a higher order p-factor. P-factor distribution within our final sample is presented in SI Figure 2.

### MRI acquisition, quality control, preprocessing and scanner harmonization

MRI data was attained on 29 different 3T scanners from Siemens Prisma, General Electric (GE) 750 and Philips. The T1w image was an inversion prepared RF-spoiled gradient echo scan, using prospective motion correction when available, and with 1 mm isotropic voxel resolution. Detailed descriptions of acquisition parameters, and care and safety procedures implemented for scanning of children are presented elsewhere (Casey et al., 2018).

We relied on the T1w quality control from the ABCD Data Analysis and Informatics Core which uses a standardized pipeline of automated and manual procedures (Hagler et al., 2019). It yields a binary code for images recommended for inclusion, and 367 youths did not pass ABCD QC and were therefore excluded. Post FreeSurfer processing (see below) we additionally excluded 104 subjects due to missing surface-based data, and 64 subjects due to total amount of surface holes being => 200 before correction (Elyounssi et al., 2023; Rosen et al., 2018).

Quality approved T1w images were processed using the open-source neuroimaging toolkit FreeSurfer 7.1 (http://surfer.nmr.mgh.harvard.edu). FreeSurfer performs volumetric segmentations and cortical surface reconstructions, including the “white” and “pial” surface, which is the grey/white matter boundary and the grey/cerebrospinal fluid (CSF) boundary, respectively (Dale et al., 1999; Fischl et al., 1999). The computation of each imaging metric including GWC is described in detail within SI. Within the current study lower GWC reflects more similar grey and white matter, a blurring that is documented across youth development (Norbom et al., 2019). Thickness, area, curvature and GWC surface maps were registered to fsaverage and smoothed using a Gaussian kernel of 15mm full width at half maximum (FWHM).

To adjust for systematic and unwanted scanner related variance, smoothed surfaces were subsequently imported to R and the package neuroCombat (Fortin et al., 2018) was employed at vertex level to harmonize data across scanners. We included five covariates to our Combat model, namely age, sex, p-factor, parental income, and parental education, to preserve such variance during the harmonization procedure. Box plots of mean or total MRI measures pre- and post-neuroCombat adjustments are presented in SI Figure 3-6.

### Multimodal fusion

Combat corrected vertex-wise surfaces of cortical thickness, surface area, curvature and GWC were fused by FMRIB’s Linked Independent Component Analysis (FLICA) (Groves et al., 2011). FLICA decomposes data into spatially independent components of variation and is robust to inputs of differing units, smoothness, and signal- and contrast to noise ratios. It can discover both multimodal features, and detect single-modality structured components if present (Groves et al., 2011, 2012). The FLICA mixing matrix vectors are statistically independent but not required to be orthogonal and can therefore account for shared variance from variables external to the FLICA. FLICA was employed with 1000 iterations, and a log-transform of surface area only. A model order of 70 was chosen based on having the highest cophenetic correlation coefficient after testing model orders ranging from 50-80. This range was chosen to balance coherent statistical analyses and interpretable findings with the ability to discern distinct patterns. The cophenetic correlation coefficient is an indication of how well the similarities of the clustering result matches subject resemblances within the original dataset.

### Statistical analyses

All demographic and behavioral data (see Figure 1 for a correlation matrix) were z-standardized, and associations between parental SES and LICA component subject loadings were tested using linear mixed effects (LME) models in R. We used the “lme4” (Bates et al., 2015), and the “lme.dscore” package from EMAtools, the latter for Cohens D calculations.

**Figure 1.**
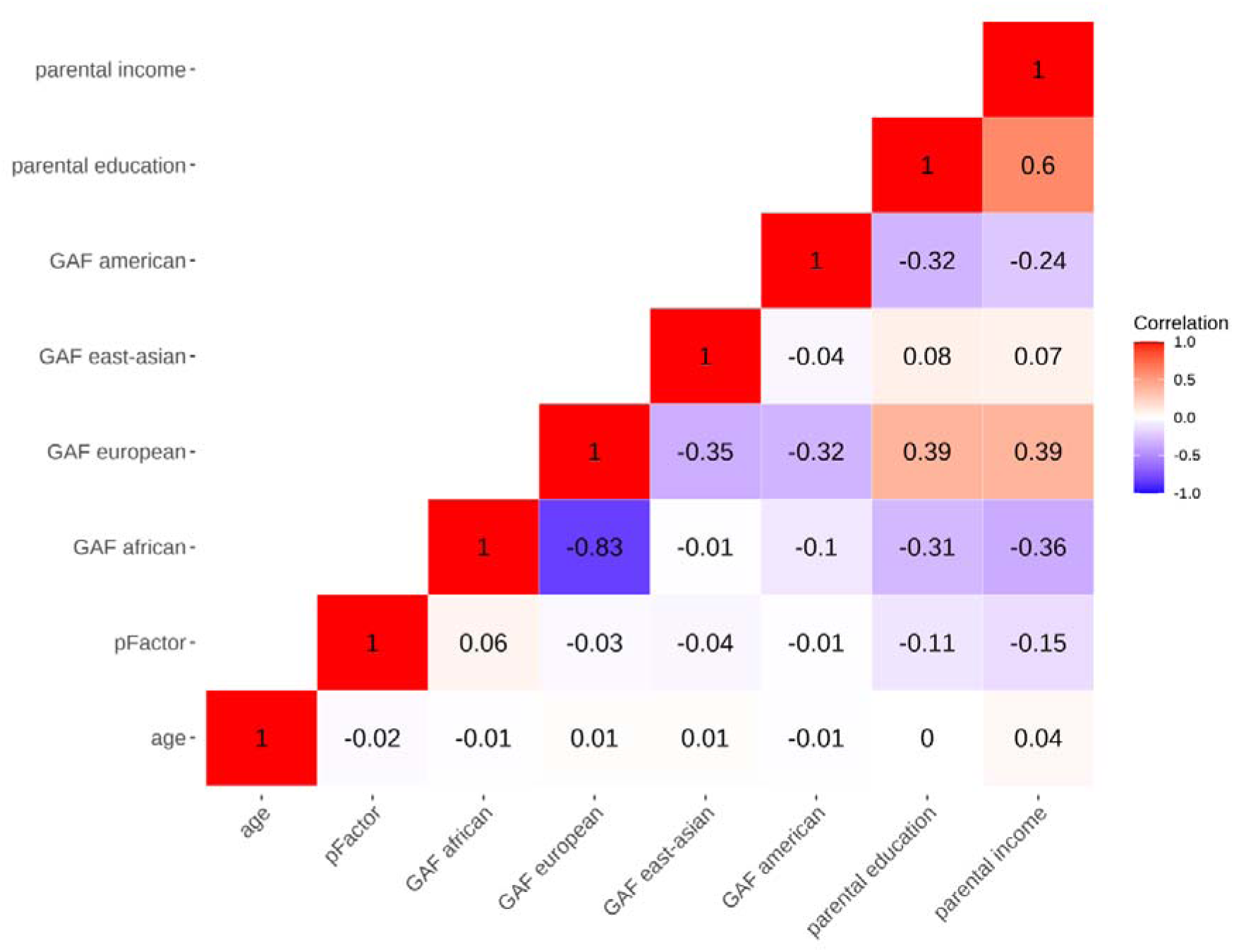
Pearson’s correlations of key demographic and behavioral variables. The figure shows a correlation matrix of all the demographic and behavioral variables from the final sample (n=9758), which were added as fixed effects and dependent variables for our main analyses.

First, we tested the linear association between parental education and the 70 LICA components in separate models. Parental education and subject loading were added as independent and dependent variables, respectively. Age, sex, and four genetic ancestry factors (GAFs) were included as fixed effects, the latter based on population inference from genetic variants (Huang et al., 2022), and were included to minimize confounds from population stratification. To maximize generalizability and statistical power, we included family ID and monozygotic twin status as random effects, to model shared environmental and genetic influences. We then ran identical analyses testing the linear association between parental income and each LICA component. Tests of the quadratic relationship between both parental SES metrics and LICA components are presented in the SI.

Second, we tested the linear association between parental SES and child p-factor scores. We included parental education or income as independent variables in separate models and p-factor scores as the dependent variable. Fixed and random effects were identical to our previous models.

Third, we tested whether relevant LICA components (defined as showing a significant association with parental SES), could moderate the relationship between parental SES and child p-factor scores. The interaction term between parental education and subject loading was included as an independent variable, while child p-factor was added as a dependent variable. Parental education, subject loading, age, sex, and GAFs were added as fixed effects, while family ID and monozygotic twin status were added as random effects. We then ran identical analyses where parental education was replaced with parental income.

Finally, to compare our multimodal results to more conventional unimodal approaches, we tested the linear association between parental SES and standardized mean cortical thickness, total surface area, mean curvature, and mean GWC in separate models.

For all statistical analyses with multiple comparisons, p-values were adjusted by false discovery rate (FDR) using Benjamini-Hochberg’s procedure and a significance threshold of 0.05. The statistical code used in the present paper can be found online (https://osf.io/etsjx/).

## Results

### Multimodal decomposition

FLICA decomposition resulted in 70 independent components (ICs) of youth cortical brain structure. Modal weighting was dominated by surface area followed by GWC, cortical thickness, and with relatively little contribution from curvature, as visualized in Figure 2. Of note, IC59, the only component showing a strong curvature weighting was highly driven by a single subject and was therefore disregarded. ICs were ordered based on total explained variance with IC1 and IC2 explaining 18.7% and 16% respectively. Both ICs were mainly unimodal, the first highly dominated by global surface area and the second by global GWC. The remaining components explained between 3.23-0.47% of total variance in the cortical decomposition, and all components should be interpreted as showing patterns beyond, or “in addition” to the other components.

**Figure 2.**
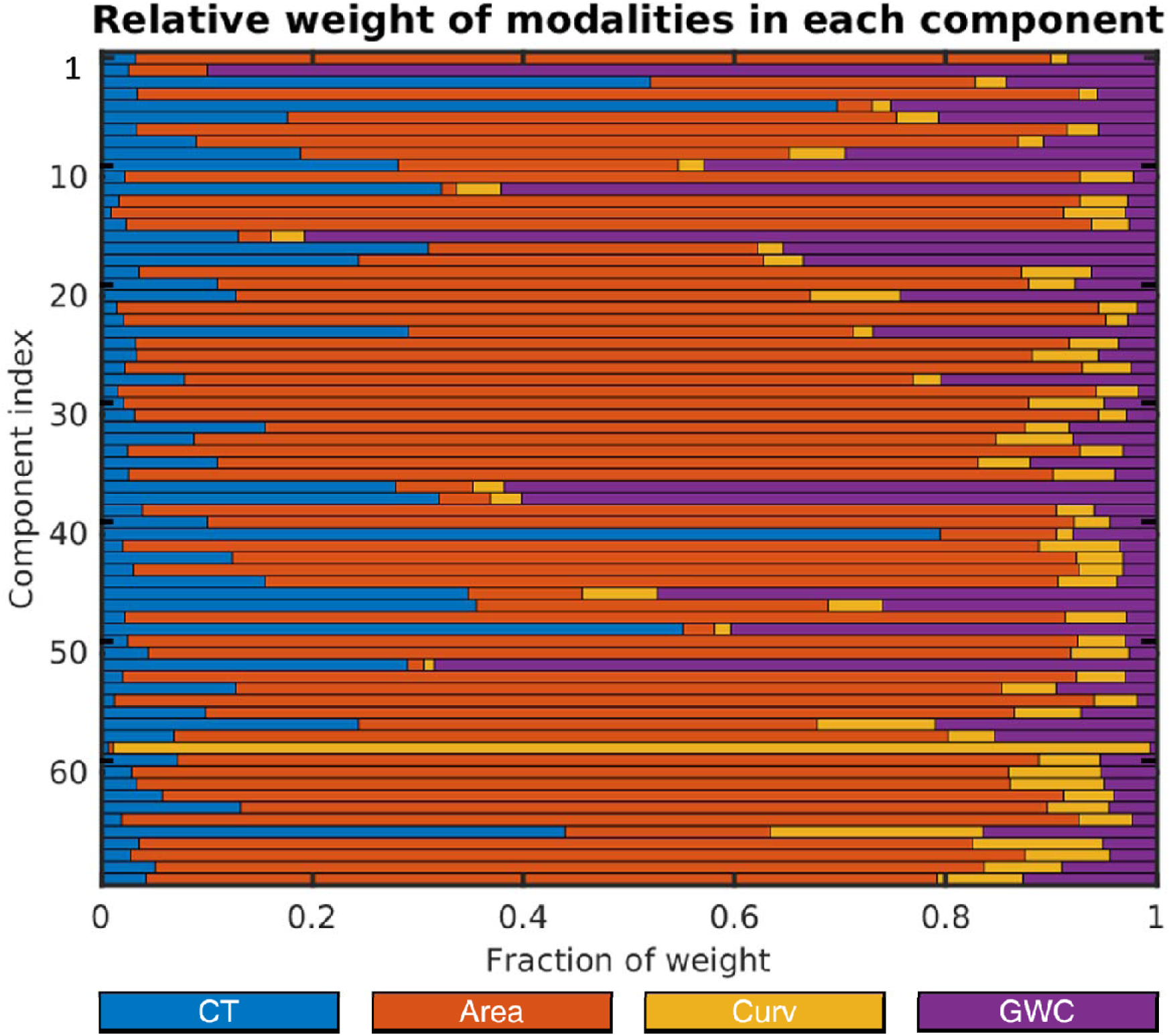
Linked independent component analysis (LICA) decomposition. The figure shows the color-coded relative weight of cortical thickness (CT), surface area (Area), curvature (Curv) and grey/white-matter contrast (GWC), within each of the 70 components.

### Linear associations between parental education and youth cortical structure

LME models revealed significant associations between parental education and six ICs of youth cortical structure, namely IC1, IC2, IC10, IC20, IC49 and IC54 (see Table 1 and Figure 3).

**Table 1.** Associations between parental socioeconomic status (SES) and cortical component loadings. The table depicts Cohens D, T statistic, and uncorrected- and FDR-corrected p-values from the statistical analyses of the associations between parental education, parental income, and each component loading. Significant p-values are marked with Asterix (*).

| IC | PARENTAL EDUCATION |  |  |  | PARENTAL INCOME |  |  |  |
| --- | --- | --- | --- | --- | --- | --- | --- | --- |
|  | Cohens D | T statistic | p-value | p-value | Cohens D | T statistic | p-value | p-value |
|  |  |  | (uncorrected) | (corrected) |  |  | (uncorrected) | (corrected) |
| 1 | 0.1 | 4.52 | <0.000* | <0.000* | 0.09 | 4.25 | <0.000* | 0.001* |
| 2 | 0.13 | 5.8 | <0.000* | <0.000* | 0.15 | 6.72 | <0.000* | <0.000* |
| 3 | 0.06 | 2.7 | 0.007* | 0.411 | 0.05 | 2.4 | 0.016* | 0.952 |
| 4 | -0.07 | -3 | 0.003* | 0.167 | -0.09 | -3.81 | <0.000* | 0.009* |
| 5 | 0 | -0.07 | 0.947 | 0.997 | -0.03 | -1.32 | 0.186 | 0.974 |
| 6 | -0.01 | -0.49 | 0.628 | 0.997 | 0 | -0.13 | 0.896 | 0.974 |
| 7 | -0.07 | -3.24 | 0.001* | 0.074 | -0.03 | -1.42 | 0.156 | 0.974 |
| 8 | 0 | -0.02 | 0.982 | 0.997 | 0.02 | 0.78 | 0.434 | 0.974 |
| 9 | 0 | 0 | 0.997 | 0.997 | 0.03 | 1.33 | 0.185 | 0.974 |
| 10 | 0.14 | 6.17 | <0.000* | <0.000* | 0.11 | 5.08 | <0.000* | <0.000* |
| 11 | 0.01 | 0.38 | 0.704 | 0.997 | -0.01 | -0.26 | 0.794 | 0.974 |
| 12 | -0.02 | -0.9 | 0.37 | 0.997 | 0.01 | 0.33 | 0.738 | 0.974 |
| 13 | 0.03 | 1.38 | 0.169 | 0.997 | 0.02 | 0.86 | 0.388 | 0.974 |
| 14 | -0.03 | -1.2 | 0.23 | 0.997 | -0.01 | -0.52 | 0.606 | 0.974 |
| 15 | -0.04 | -1.6 | 0.11 | 0.997 | -0.04 | -1.56 | 0.118 | 0.974 |
| 16 | 0.02 | 1.06 | 0.291 | 0.997 | 0.01 | 0.62 | 0.537 | 0.974 |
| 17 | -0.03 | -1.49 | 0.136 | 0.997 | -0.05 | -2.35 | 0.019* | 0.974 |
| 18 | -0.01 | -0.64 | 0.522 | 0.997 | -0.01 | -0.63 | 0.528 | 0.974 |
| 19 | -0.01 | -0.47 | 0.635 | 0.997 | -0.05 | -2.12 | 0.034* | 0.974 |
| 20 | 0.09 | 4.13 | <0.000* | 0.002* | 0.08 | 3.8 | <0.000* | 0.01* |
| 21 | 0.04 | 2.02 | 0.044* | 0.997 | 0.07 | 3.23 | 0.001* | 0.077 |
| 22 | 0.05 | 2.42 | 0.016* | 0.899 | 0.02 | 0.93 | 0.352 | 0.974 |
| 23 | -0.01 | -0.34 | 0.732 | 0.997 | 0.01 | 0.35 | 0.729 | 0.974 |
| 24 | 0.02 | 0.95 | 0.34 | 0.997 | 0.02 | 1.01 | 0.311 | 0.974 |
| 25 | -0.06 | -2.64 | 0.008* | 0.486 | -0.05 | -2.3 | 0.021* | 0.974 |
| 26 | -0.04 | -1.62 | 0.106 | 0.997 | -0.03 | -1.26 | 0.207 | 0.974 |
| 27 | 0 | 0.07 | 0.946 | 0.997 | -0.03 | -1.49 | 0.136 | 0.974 |
| 28 | -0.02 | -0.7 | 0.483 | 0.997 | -0.03 | -1.36 | 0.175 | 0.974 |
| 29 | -0.05 | -2.22 | 0.027* | 0.997 | 0 | 0.17 | 0.864 | 0.974 |
| 30 | -0.02 | -0.68 | 0.495 | 0.997 | -0.02 | -0.97 | 0.331 | 0.974 |
| 31 | 0.04 | 1.61 | 0.108 | 0.997 | 0 | 0.15 | 0.882 | 0.974 |
| 32 | 0.01 | 0.27 | 0.791 | 0.997 | 0.03 | 1.2 | 0.23 | 0.974 |
| 33 | 0.04 | 1.77 | 0.076 | 0.997 | 0 | -0.05 | 0.964 | 0.974 |
| 34 | 0.01 | 0.41 | 0.681 | 0.997 | 0.01 | 0.43 | 0.671 | 0.974 |
| 35 | -0.01 | -0.29 | 0.775 | 0.997 | -0.02 | -0.72 | 0.472 | 0.974 |
| 36 | -0.01 | -0.25 | 0.8 | 0.997 | 0.01 | 0.5 | 0.618 | 0.974 |
| 37 | -0.03 | -1.42 | 0.156 | 0.997 | -0.01 | -0.56 | 0.579 | 0.974 |
| 38 | 0 | -0.17 | 0.866 | 0.997 | -0.02 | -1.05 | 0.295 | 0.974 |
| 39 | -0.04 | -1.97 | 0.049* | 0.997 | -0.02 | -0.69 | 0.492 | 0.974 |
| 40 | -0.01 | -0.36 | 0.718 | 0.997 | 0 | -0.22 | 0.828 | 0.974 |
| 41 | -0.06 | -2.8 | 0.005* | 0.313 | -0.03 | -1.31 | 0.191 | 0.974 |
| 42 | 0 | -0.16 | 0.872 | 0.997 | -0.01 | -0.49 | 0.624 | 0.974 |
| 43 | -0.03 | -1.31 | 0.191 | 0.997 | -0.05 | -2.24 | 0.025 | 0.974 |
| 44 | -0.03 | -1.49 | 0.136 | 0.997 | 0 | 0.03 | 0.974 | 0.974 |
| 45 | -0.04 | -1.82 | 0.069 | 0.997 | -0.07 | -3.28 | 0.001* | 0.065 |
| 46 | -0.03 | -1.31 | 0.192 | 0.997 | 0.04 | 1.88 | 0.061 | 0.974 |
| 47 | 0 | -0.06 | 0.952 | 0.997 | 0.05 | 2.07 | 0.039* | 0.974 |
| 48 | -0.04 | -1.64 | 0.101 | 0.997 | -0.03 | -1.3 | 0.194 | 0.974 |
| 49 | 0.09 | 4.17 | <0.000* | 0.002* | 0.1 | 4.3 | <0.000* | 0.001 |
| 50 | 0.03 | 1.33 | 0.184 | 0.997 | -0.02 | -1.07 | 0.286 | 0.974 |
| 51 | 0.02 | 0.88 | 0.381 | 0.997 | 0.04 | 1.53 | 0.126 | 0.974 |
| 52 | -0.03 | -1.46 | 0.144 | 0.997 | 0 | -0.1 | 0.92 | 0.974 |
| 53 | 0.01 | 0.63 | 0.53 | 0.997 | 0.04 | 1.7 | 0.09 | 0.974 |
| 54 | -0.09 | -4.13 | <0.000* | 0.002 | -0.08 | -3.41 | 0.001* | 0.041* |
| 55 | 0.04 | 1.84 | 0.066 | 0.997 | 0.06 | 2.47 | 0.013* | 0.791 |
| 56 | 0 | 0.18 | 0.856 | 0.997 | 0.01 | 0.65 | 0.516 | 0.974 |
| 57 | 0 | -0.16 | 0.871 | 0.997 | -0.01 | -0.33 | 0.743 | 0.974 |
| 58 | 0.03 | 1.23 | 0.218 | 0.997 | 0.04 | 1.67 | 0.095 | 0.974 |
| 59 | -0.33 | - | - | - | 0.17 | - | - | - |
| 60 | 0.05 | 2.25 | 0.025* | 0.997 | 0.02 | 1.07 | 0.286 | 0.974 |
| 61 | -0.03 | -1.24 | 0.214 | 0.997 | -0.01 | -0.33 | 0.738 | 0.974 |
| 62 | 0.01 | 0.39 | 0.696 | 0.997 | -0.03 | -1.29 | 0.197 | 0.974 |
| 63 | 0.03 | 1.31 | 0.192 | 0.997 | 0.02 | 0.96 | 0.335 | 0.974 |
| 64 | -0.04 | -1.73 | 0.085 | 0.997 | -0.06 | -2.53 | 0.012* | 0.694 |
| 65 | -0.02 | -0.73 | 0.465 | 0.997 | -0.03 | -1.23 | 0.219 | 0.974 |
| 66 | -0.03 | -1.42 | 0.156 | 0.997 | -0.05 | -2.26 | 0.024* | 0.974 |
| 67 | 0 | 0.19 | 0.853 | 0.997 | -0.04 | -1.66 | 0.098 | 0.974 |
| 68 | 0.03 | 1.15 | 0.251 | 0.997 | 0.04 | 1.64 | 0.102 | 0.974 |
| 69 | -0.01 | -0.51 | 0.608 | 0.997 | 0 | -0.18 | 0.855 | 0.974 |
| 70 | 0.02 | 0.86 | 0.392 | 0.997 | 0.03 | 1.38 | 0.166 | 0.974 |

**Figure 3.**
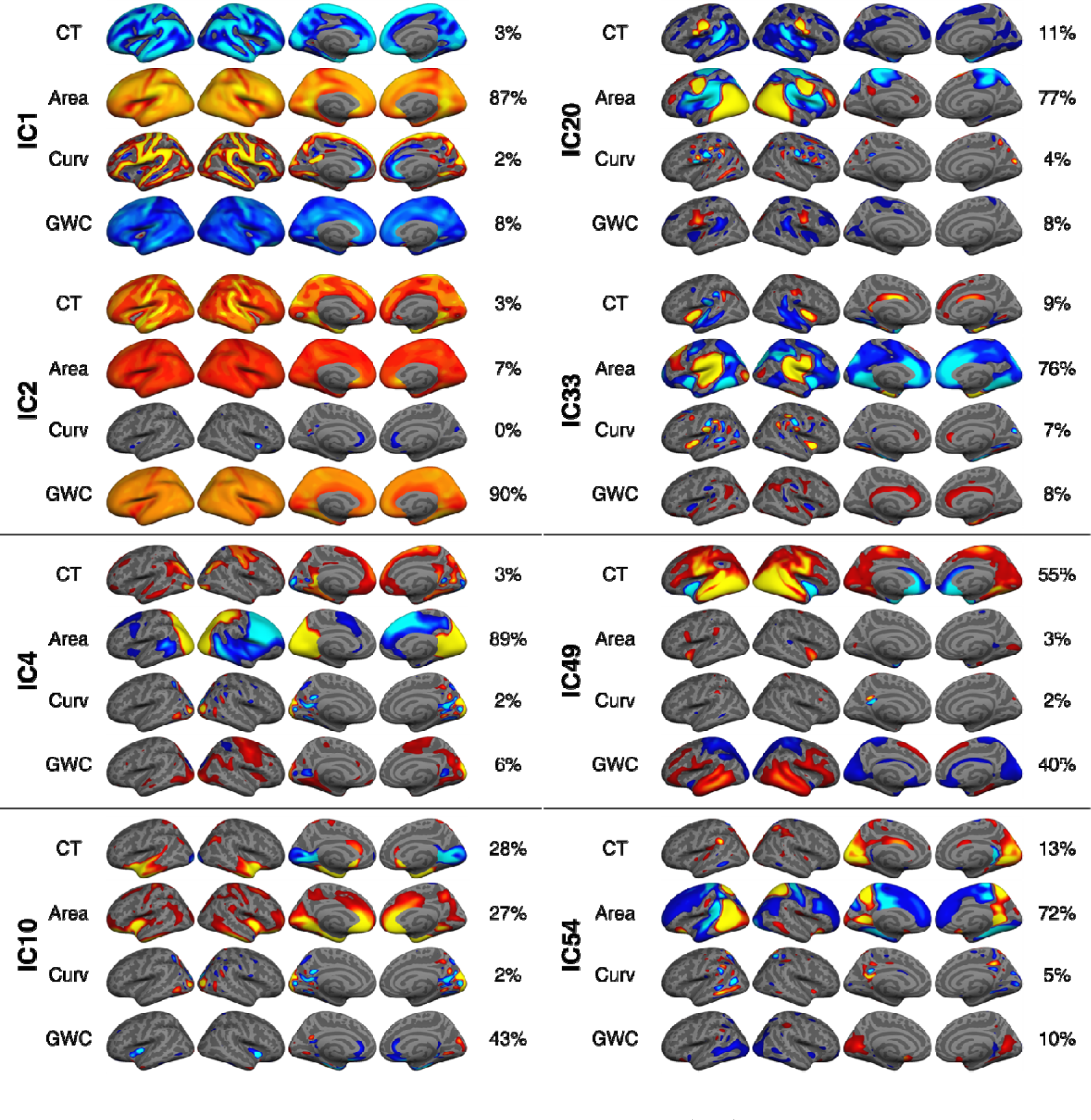
Spatial visualization of independent components (ICs) showing a significant association with parental education and/or income. All components were thresholded with a minimum and maximum of 7 and 20 standard deviations, respectively, except for surface area within IC1 and IC2, and GWC within IC2, which were thresholded with a higher value to reveal nuances in the global pattern.

Parental education showed a positive association with IC1, which was a bi-hemispheric component, dominated by larger global surface area (87%) and to a lesser extent lower global GWC (8%). This indicates that parental education is associated with larger cortical surface area and lower GWC in child offspring.

Parental education showed a stronger positive association with IC2 than with IC1. IC2 was a bi-hemispheric and mostly unimodal component of higher global GWC (90%) and to a lesser extent larger surface area (7%). Relative to the findings of IC1, this indicates that parental education is somewhat linked to higher GWC globally and larger surface area in childhood.

The strongest association for parental education was a positive association with IC10. IC10 was bi-hemispheric and multimodal, capturing a joint pattern of lower GWC (43%) and higher thickness (28%) and surface area (27%) within temporal pole and insular regions, as well as lower medial occipital thickness. This indicates that beyond the global findings already described, parental education is associated with lower GWC, thicker cortex and larger surface area in insular cortical regions in childhood.

Parental education showed a positive association with IC20, which showed a bi-hemispheric pattern of larger occipital and smaller parietal surface area (77%) as well as an overlapping bidirectional and local thickness (11%) and GWC (8%) pattern. This indicates that parental education is linked to additional local variations in child cortical structure, particularly of larger surface area.

Parental education showed a positive association with IC49, a component with a bi-hemispheric pattern of higher thickness (55%) within occipital, temporal and parietal regions extending into the frontal lobe, as well as a bi-directional pattern for GWC in similar regions (40%). Parental education is therefore linked to additional local variations, particularly of higher cortical thickness and higher and lower GWC locally in childhood.

Parental education showed a negative association with IC54. It generally showed a bi-hemispheric pattern of larger occipital and smaller frontal surface area (77%), larger medial occipital thickness (13%), and higher medial occipital-, and lower lateral occipital GWC (10%). This suggests that parental education is linked to further local variations in child cortical structure, particularly of smaller occipital and larger frontal surface area.

In summary, our results suggest that children with more educated parents have higher GWC and larger surface area globally, as well as additional local cortical variations of larger frontal and insular surface area and to a lesser extent bidirectional GWC patterns and higher thickness.

### Linear associations between parental income and youth cortical structure

LME models revealed significant associations between parental income and 7 ICs of youth cortical structure, namely IC1, IC2, IC4, IC10, IC20, IC49 and IC54 (see Table 1 and Figure 3).

In addition to the ICs already described above, parental income showed a negative association with IC4. IC4 was dominated by larger occipito-parietal- and smaller frontal surface area (89%) of the right hemisphere, and to a lesser extent higher right hemisphere frontal and occipital GWC (6%). This indicates that beyond the relations already described, parental income is associated with local, right-hemisphere variations in child cortical structure, of smaller occipito-parietal and larger frontal surface area, as well as lower local GWC to a lesser extent. Quadratic associations between parental income and LICA components are presented in the SI and SI Table 2. The spread of effect sizes for ICs significantly associated with SES subfactors across scanners are presented in SI Figure 8.

### Linear associations between parental SES and child p-factor, and moderation effects of cortical structure

LME models revealed significant negative associations between the child p-factor scores and both parental education (*d*=-0.24, t=-10.90, *p*= <.001) and parental income (*d*=-0.31, t=-13.67, *p*= <.001). We then tested whether the negative parental SES – p-factor relations were moderated by individual differences in cortical structure.

LME models revealed no moderation effect of cortical structure on the parental education - child p-factor relationship (SI Table 3). We found a significant negative moderation effect of IC54 (*d*= -0.07, t= -3.06, corrected p= 0.016) on the negative relationship between parental income and child p-factor scores (Figure 4 and SI Table 3). This indicates that the relationship between parental income and psychopathology in childhood is attenuated for children with larger frontal-, and smaller occipital surface area and lower medial occipital thickness and GWC.

**Figure 4.**
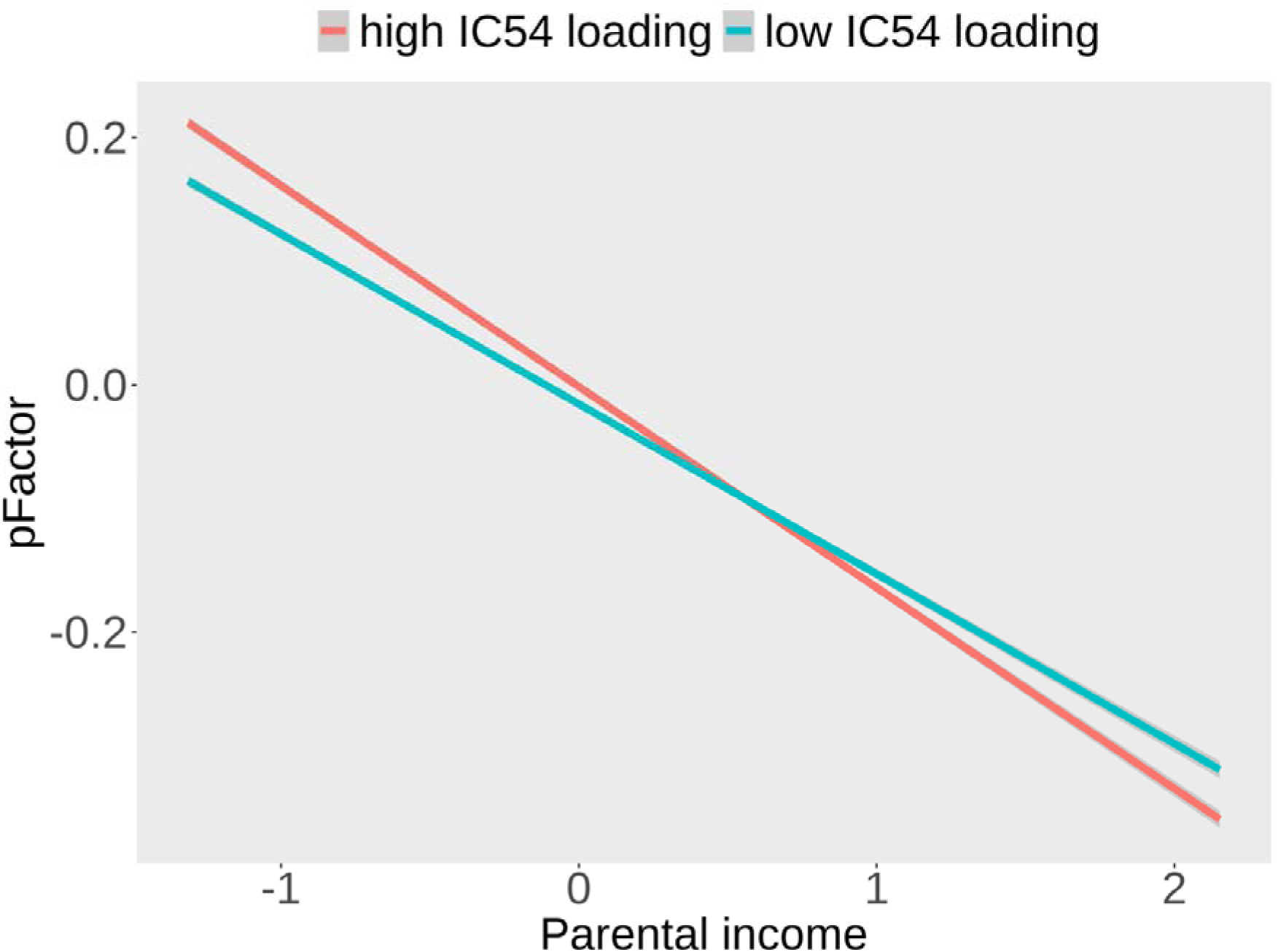
A visualization of the moderation effect of IC54 on the relationship between parental income and child p-factor scores. For visualization, the sample is divided by median split, so that subjects who show a high loading on IC54 are marked in red, and subjects who show a low loading on IC54 are marked in green.

### Linear associations between parental SES and unimodal global cortical metrics

To compare our findings to standard unimodal assessments, we ran LME models testing the association between parental SES and unimodal metrics, separately. These results are described in the SI and SI Table 4. In short, unimodal assessments showed close correspondence to our multimodal fusion analyses, with significant positive associations for both parental education and parental income and total surface area as well as mean GWC. We did not find any significant associations between either parental SES metric and mean cortical thickness or mean curvature.

## Discussion

We sought to characterize the relationships between the socioeconomic environment and a child’s cortical structure, and whether it moderated the association between parental SES and child psychopathology. Children from more educated and affluent parents had a combination of larger surface area and GWC, in addition to local variations of larger surface area and to a lesser extent bidirectional GWC and thickness patterns. The negative relationship between parental income and child psychopathology was attenuated for children showing a pattern of larger frontal and smaller occipital surface area and lower medial occipital thickness and GWC.

The multimodal decomposition of vertex-wise cortical metrics revealed that most of the variance within the data could be explained by global surface area, and global GWC. Despite the participants’ relatively young age, the prominence of global surface area could in part be due to sex-related differences. As expected in a sample with a narrow age range, cortical thickness was less dominant, as was curvature with minimal contribution. A more detailed discussion of our FLICA decomposition is discussed within the SI.

In line with previous studies (Noble et al., 2015; Rakesh & Whittle, 2021) our results showed that parental education and income were both associated with larger cortical surface area in children. The neurobiology underlying MRI based area differences in late childhood are not fully understood. Increased pericortical myelin and axon calibers could possibly be protruding- and pushing the cortical surface outward, increasing its size, while at the same time decreasing cortical folding and thickness (Seldon, 2005). Indeed, although contributions were small, fused cortical thickness and GWC showed global negative patterns in accordance with a thinner and brighter cortex. Curvature, on the other hand, did not show this pattern.

Children from higher SES families also showed globally higher GWC, emerging as the strongest finding for parental income and, apart from a multimodal insular pattern, also for parental education. While it can be challenging to interpret separate components with opposing patterns, our unimodal analyses concordantly showed a positive association between both parental SES metrics and global GWC. There has to our knowledge been no previous tests of the relations between parental SES and child GWC. However, our findings correspond well with a recent paper that assessed parental SES and its associations to the related, but directionally inverse, intensity metric T1w/T2w ratio (Norbom et al., 2022). Here, widespread negative relations were found between general- and subfactor parental SES and T1w/T2w ratio. Effect sizes were also larger than for standard morphometry, including cortical surface area and cortical thickness. GWC thus appears to be a highly sensitive imaging marker not only for youth cortical development generally (Norbom et al., 2019, 2020), but also specifically for SES related variance.

From a neurobiological standpoint, although consistently reported and thus in line with our hypothesis, the direction of GWC results is counterintuitive. GWC decreases across childhood and adolescence, pointing to grey and white matter intensities becoming more similar, possibly due to higher levels of intracortical myelin. Our results may thus indicate that children from lower SES families have a more developed cortex. Possible reasons for the direction of findings are discussed in detail elsewhere (Norbom et al., 2022), but include a disadvantageous effect of excess myelin, cortical surface misclassification, and other tissue properties accounting for GWC variations. On the other hand, our results fit with the theory that exposure to poverty and early adversity can foster accelerated maturation (Belsky, 2019; Colich et al., 2020). This notion has also been supported by MRI based studies (Colich et al., 2020). It would be beneficial to employ quantitative relaxometry to further probe the underlying neurobiology linking SES to GWC.

Multimodal fusion revealed that parental education and income were associated with complex multimodal cortical patterns, including larger frontal and insular surface area, coupled with lower insular GWC and thickness. Although previous research is discrepant, the most consistent morphometric pattern is indeed a positive association between parental SES and frontal surface area (Rakesh & Whittle, 2021). Although based on large tissue volumes, overlapping multimodal patterns should improve our inferences of the underlying neurobiology. For instance, children from higher-income parents had right hemisphere increases in frontal surface area weakly coupled with thinner cortex and lower GWC. Similarly, our insular component coupled larger surface area with lower GWC. In youth MRI studies, these are well documented maturational patterns (Norbom et al., 2021) that in sum are understood to reflect a combination of pericortical and cortical myelination, increased axon caliber, remodeling of dendritic arbour and reductions in glial cells (Huttenlocher & Dabholkar, 1997; Petanjek et al., 2008, 2011; Peter R., 1979; Seldon, 2005; Vidal-Pineiro et al., 2020). While structure to function relations are not analogous, future fMRI studies could assess whether the socioeconomic environment also affects frontal cortex and insular function.

As expected, parental SES was not closely linked to cortical thickness in youth, only showing a few highly local variations within multimodal components. Findings were corroborated by unimodal mean thickness analyses showing no significant associations to either parental SES metric. This is in line with previous research pointing to mixed- and mostly null findings regarding parental SES and cortical thickness (Rakesh & Whittle, 2021). Yet, our observations underscore the advantage of multimodal fusion in discerning subtle patterns that are not captured in unimodal thickness assessments. Overall, our findings could indicate that cortical thinning across youth might be affected more by genetic factors, while surface area (Strike et al., 2019) and GWC could be more sensitive to environmental impact. However, a recent ABCD study found a relation between family income and ROI based cortical thickness (Tomasi & Volkow, 2021), as did several papers testing the related concept of neighborhood disadvantage (Rakesh & Whittle, 2021; Taylor et al., 2020; Vargas et al., 2020). A similar study by Hackman et al. (2021) did not corroborate these findings.

Multi- and unimodal assessments did not reveal an association between child socioeconomic environment and cortical curvature. Developmental folding differences across youth have been less explored, and to our knowledge no studies have previously assessed the SES - youth curvature relation. Nevertheless, a multivariate brain structure–behavior mapping revealed several modes of covariation, including one capturing general economic deprivation, and the related metric sulcal depth (Alnæs et al., 2020).

In line with our hypothesis and current literature (Letourneau et al., 2013; Reiss, 2013) children from less educated and lower income families were reported to have higher levels of general psychopathology. We tested whether cortical macro-, and microstructure moderate this relationship. We found a small moderation indicating that in low-income families child psychopathology were attenuated in children with a multimodal pattern of larger frontal- and smaller occipital surface area, and lower medial occipital thickness and GWC. Within higher income families where general psychopathology levels are low, we found the reverse pattern. Still, this finding should be interpreted with caution awaiting replication. Future studies should also employ longitudinal designs to test whether cortical structure mediates the relation between parental SES and future psychopathology in youth (Farah, 2017; Maxwell & Cole, 2007).

There are several limitations to our study. First, regarding our SES estimations, we did not include subjective or cultural aspects of the construct (W. M. Liu et al., 2004; Yosso, 2005) nor did we investigate the unique contributions of our SES metrics. Also, income and education are distal markers for proximal causal pathways including stress, cognitive stimulation, obstetric complications, prenatal care, toxins and nutrition (Evans & Kim, 2013; Farah, 2017; Thomas & Coecke, 2023). Second, our study design can only capture correlational relationships and cannot infer causation. Similarly, longitudinal MRI data is needed to test the maturation of cortical structure and its relation to SES over time. Third, several brain related findings including the moderation effect, were of small magnitude. This is not surprising as publication bias and historically small samples may have promoted inflated neurodevelopmental effects (Button et al., 2013; Ioannidis, 2008) that should be attenuated when assessing ABCD (Dick et al., 2021). Also, for many outcomes in nature causal relations are in reality small (Dick et al., 2021), including clinical effects within psychology and psychiatry (Meyer et al., 2001), and in multimodal imaging and health outcomes (Miller et al., 2016).

It is presently unclear *how* childhood socioeconomic diversity becomes neurobiologically embedded and influence current and future risk for mental health problems. In the present study we report that children with more educated or affluent parents have a combination of larger global and regional surface area, a larger difference between grey and white matter intensities, and that in low-income families, having what appears to be a more developed cortex across metrics is beneficial to mental health.

## Data Availability

All ABCD data is stored in the NIMH Data Archive Collection #2573, which is available for registered and authorised users (Request #7474, PI: Westlye). The 4.0 release will be permanently available as a persistent dataset defined in the NDA Study 1299 and has been assigned the DOI 10.15154/1523041

## Acknowledgments

Data was obtained from the Adolescent Brain Cognitive Development (ABCD) Study (https://abcdstudy.org), wich is supported by the National Institutes of Health and additional federal partners under award numbers: U01DA041048,U01DA050989,U01DA051016,U01DA041022,U01DA051018,U01DA0510 37,U01DA050987,U01DA041174,U01DA041106,U01DA041117,U01DA041028,U01DA04 1134,U01DA050988,U01DA051039,U01DA041156,U01DA041025,U01DA041120,U01DA 051038,U01DA041148,U01DA041093,U01DA041089,U24DA041123,U24DA041147. A full list of supporters is available at https://abcdstudy.org/federal-partners.html. A listing of participating sites and study investigators can be found at https://abcdstudy.org/consortium_members/. ABCD consortium investigators designed and implemented the study and/or provided data but did not necessarily participate in the analysis or writing of this report. This manuscript reflects the views of the authors and may not reflect the opinions or views of the NIH or ABCD consortium investigators.

Funding: This work was supported by the Research Council of Norway (#223273,#288083, #323951,#249795,#248238,#276082,#286838,#288083), the South-Eastern Norway Regional Health Authority (#2019069,#2021070,#2023012,#500189,#2014097,#2015044,#2015073,#2016083, #2018037,#2018076, #2019101), KG Jebsen Stiftelsen, the ERA-Net Cofund through the ERA PerMed project IMPLEMENT, and the European Research Council under the European Union’s Horizon 2020 research and Innovation program (ERC StG Grant #802998)

## Notes

### Competing Interest Statement

The authors have declared no competing interest.

### Funding Statement

This work was supported by the Research Council of Norway (#223273, #288083, #323951, #249795, #248238, #276082, #286838, and #288083), the South-Eastern Norway Regional Health Authority (#2019069, #2021070, #2023012, #500189, #2014097, #2015044, #2015073, #2016083, #2018037, #2018076, and #2019101), KG Jebsen Stiftelsen, the ERA-Net Cofund through the ERA PerMed project IMPLEMENT, and the European Research Council under the European Unions Horizon 2020 research and Innovation program (ERC StG Grant #802998).

### Author Declarations

The Institutional Review Board at the University of California, San Diego, approved all aspects of ABCD Study. The current study was conducted in line with the Declaration of Helsinki and was approved by the Regional Committee for Medical and Health Research Ethics (REK 2019/943)

### Summary of Updates

Supplementary Figure 8 Motivation for choosing our SES subfactors A link to code used in our statistical analyses

## References

1. Achenbach, T. M., & Ruffle, T. M. (2000). The Child Behavior Checklist and Related Forms for Assessing Behavioral/Emotional Problems and Competencies. Pediatrics In Review, 21(8), 265–271. 10.1542/pir.21.8.265

2. Alnæs, D., Kaufmann, T., Marquand, A. F., Smith, S. M., & Westlye, L. T. (2020). Patterns of sociocognitive stratification and perinatal risk in the child brain. Proceedings of the National Academy of Sciences, 117(22), 12419–12427. 10.1073/pnas.2001517117

3. Bartzokis, G. (2012). Neuroglialpharmacology: Myelination as a shared mechanism of action of psychotropic treatments. Neuropharmacology, 62(7), 2137–2153. 10.1016/j.neuropharm.2012.01.015

4. Bates, D., Mächler, M., Bolker, B., & Walker, S. (2015). Fitting Linear Mixed-Effects Models Using lme4. Journal of Statistical Software, 67(1). 10.18637/jss.v067.i01

5. Baumann, N., & Pham-Dinh, D. (2001). Biology of Oligodendrocyte and Myelin in the Mammalian Central Nervous System. Physiological Reviews, 81(2), 871–927. 10.1152/physrev.2001.81.2.871

6. Belsky, J. (2019). Early-Life Adversity Accelerates Child and Adolescent Development. Current Directions in Psychological Science, 28(3), 241–246. 10.1177/0963721419837670

7. Bourdieu, P. (2011). The forms of capital.(1986). Cultural Theory: An Anthology, 1(81–93), 949.

8. Button, K. S., Ioannidis, J. P. A., Mokrysz, C., Nosek, B. A., Flint, J., Robinson, E. S. J., & Munafò, M. R. (2013). Power failure: Why small sample size undermines the reliability of neuroscience. Nature Reviews Neuroscience, 14(5), 365–376. 10.1038/nrn3475

9. Casey, B. J., Cannonier, T., Conley, M. I., Cohen, A. O., Barch, D. M., Heitzeg, M. M., Soules, M. E., Teslovich, T., Dellarco, D. V., Garavan, H., Orr, C. A., Wager, T. D., Banich, M. T., Speer, N. K., Sutherland, M. T., Riedel, M. C., Dick, A. S., Bjork, J. M., Thomas, K. M., … Dale, A. M. (2018). The Adolescent Brain Cognitive Development (ABCD) study: Imaging acquisition across 21 sites. Developmental Cognitive Neuroscience, 32, 43–54. 10.1016/j.dcn.2018.03.001

10. Clark, D. A., Hicks, B. M., Angstadt, M., Rutherford, S., Taxali, A., Hyde, L., Weigard, A. S., Heitzeg, M. M., & Sripada, C. (2021). The General Factor of Psychopathology in the Adolescent Brain Cognitive Development (ABCD) Study: A Comparison of Alternative Modeling Approaches. Clinical Psychological Science, 9(2), 169–182. 10.1177/2167702620959317

11. Colich, N. L., Rosen, M. L., Williams, E. S., & McLaughlin, K. A. (2020). Biological aging in childhood and adolescence following experiences of threat and deprivation: A systematic review and meta-analysis. Psychological Bulletin, 146(9), 721–764. 10.1037/bul0000270

12. Dale, A. M., Fischl, B., & Sereno, M. I. (1999). Cortical Surface-Based Analysis. NeuroImage, 9(2), 179–194. 10.1006/nimg.1998.0395

13. Dearing, E., McCartney, K., & Taylor, B. A. (2006). Within-child associations between family income and externalizing and internalizing problems. Developmental Psychology, 42(2), 237–252. 10.1037/0012-1649.42.2.237

14. Desikan, R. S., Ségonne, F., Fischl, B., Quinn, B. T., Dickerson, B. C., Blacker, D., Buckner, R. L., Dale, A. M., Maguire, R. P., Hyman, B. T., Albert, M. S., & Killiany, R. J. (2006). An automated labeling system for subdividing the human cerebral cortex on MRI scans into gyral based regions of interest. NeuroImage, 31(3), 968–980. 10.1016/j.neuroimage.2006.01.021

15. Destrieux, C., Fischl, B., Dale, A., & Halgren, E. (2010). Automatic parcellation of human cortical gyri and sulci using standard anatomical nomenclature. NeuroImage, 53(1), 1–15. 10.1016/j.neuroimage.2010.06.010

16. Dick, A. S., Lopez, D. A., Watts, A. L., Heeringa, S., Reuter, C., Bartsch, H., Fan, C. C., Kennedy, D. N., Palmer, C., Marshall, A., Haist, F., Hawes, S., Nichols, T. E., Barch, D. M., Jernigan, T. L., Garavan, H., Grant, S., Pariyadath, V., Hoffman, E., … Thompson, W. K. (2021). Meaningful associations in the adolescent brain cognitive development study. NeuroImage, 239, 118262. 10.1016/j.neuroimage.2021.118262

17. Duyme, M., Dumaret, A.-C., & Tomkiewicz, S. (1999). How can we boost IQs of “dull children”?: A late adoption study. Proceedings of the National Academy of Sciences, 96(15), 8790–8794. 10.1073/pnas.96.15.8790

18. Elyounssi, S., Kunitoki, K., Clauss, J. A., Laurent, E., Kane, K., Hughes, D. E., Hopkinson, C. E., Bazer, O., Sussman, R. F., Doyle, A. E., Lee, H., Tervo-Clemmens, B., Eryilmaz, H., Gollub, R. L., Barch, D. M., Satterthwaite, T. D., Dowling, K. F., & Roffman, J. L. (2023). *Uncovering and mitigating bias in large, automated MRI analyses of brain development* [Preprint]. Neuroscience. 10.1101/2023.02.28.530498

19. Evans, G. W., & Kim, P. (2013). Childhood Poverty, Chronic Stress, Self-Regulation, and Coping. Child Development Perspectives, 7(1), 43–48. 10.1111/cdep.12013

20. Farah, M. J. (2017). The Neuroscience of Socioeconomic Status: Correlates, Causes, and Consequences. Neuron, 96(1), 56–71. 10.1016/j.neuron.2017.08.034

21. Feldstein Ewing, S. W., Bjork, J. M., & Luciana, M. (2018). Implications of the ABCD study for developmental neuroscience. Developmental Cognitive Neuroscience, 32, 161–164. 10.1016/j.dcn.2018.05.003

22. Fischl, B., Sereno, M. I., & Dale, A. M. (1999). Cortical Surface-Based Analysis. NeuroImage, 9(2), 195–207. 10.1006/nimg.1998.0396

23. Fortin, J.-P., Cullen, N., Sheline, Y. I., Taylor, W. D., Aselcioglu, I., Cook, P. A., Adams, P., Cooper, C., Fava, M., McGrath, P. J., McInnis, M., Phillips, M. L., Trivedi, M. H., Weissman, M. M., & Shinohara, R. T. (2018). Harmonization of cortical thickness measurements across scanners and sites. NeuroImage, 167, 104–120. 10.1016/j.neuroimage.2017.11.024

24. Groves, A. R., Beckmann, C. F., Smith, S. M., & Woolrich, M. W. (2011). Linked independent component analysis for multimodal data fusion. NeuroImage, 54(3), 2198–2217. 10.1016/j.neuroimage.2010.09.073

25. Groves, A. R., Smith, S. M., Fjell, A. M., Tamnes, C. K., Walhovd, K. B., Douaud, G., Woolrich, M. W., & Westlye, L. T. (2012). Benefits of multi-modal fusion analysis on a large-scale dataset: Life-span patterns of inter-subject variability in cortical morphometry and white matter microstructure. NeuroImage, 63(1), 365–380. 10.1016/j.neuroimage.2012.06.038

26. Hackman, D. A., Cserbik, D., Chen, J.-C., Berhane, K., Minaravesh, B., McConnell, R., & Herting, M. M. (2021). Association of Local Variation in Neighborhood Disadvantage in Metropolitan Areas With Youth Neurocognition and Brain Structure. JAMA Pediatrics, 175(8), e210426. 10.1001/jamapediatrics.2021.0426

27. Hagler, D. J., Hatton, SeanN., Cornejo, M. D., Makowski, C., Fair, D. A., Dick, A. S., Sutherland, M. T., Casey, B. J., Barch, D. M., Harms, M. P., Watts, R., Bjork, J. M., Garavan, H. P., Hilmer, L., Pung, C. J., Sicat, C. S., Kuperman, J., Bartsch, H., Xue, F., … Dale, A. M. (2019). Image processing and analysis methods for the Adolescent Brain Cognitive Development Study. NeuroImage, 202, 116091. 10.1016/j.neuroimage.2019.116091

28. Hill, W. D., Davies, N. M., Ritchie, S. J., Skene, N. G., Bryois, J., Bell, S., Di Angelantonio, E., Roberts, D. J., Xueyi, S., Davies, G., Liewald, D. C. M., Porteous, D. J., Hayward, C., Butterworth, A. S., McIntosh, A. M., Gale, C. R., & Deary, I. J. (2019). Genome-wide analysis identifies molecular systems and 149 genetic loci associated with income. Nature Communications, 10(1), 5741. 10.1038/s41467-019-13585-5

29. Hogstrom, L. J., Westlye, L. T., Walhovd, K. B., & Fjell, A. M. (2013). The Structure of the Cerebral Cortex Across Adult Life: Age-Related Patterns of Surface Area, Thickness, and Gyrification. Cerebral Cortex, 23(11), 2521–2530. 10.1093/cercor/bhs231

30. Huang, T. H., Loughnan, R., Thompson, W. K., & Fan, C. C. (2022). *The Impact of Population Stratification on the Analysis of Multimodal Neuroimaging Derived Measures* [Preprint]. Neuroscience. 10.1101/2022.08.06.503037

31. Huttenlocher, P. R., & Dabholkar, A. S. (1997). Regional differences in synaptogenesis in human cerebral cortex. The Journal of Comparative Neurology, 387(2), 167–178. 10.1002/(SICI)1096-9861(19971020)387:2<167::AID-CNE1>3.0.CO;2-Z

32. Ioannidis, J. P. A. (2008). Why Most Discovered True Associations Are Inflated. Epidemiology, 19(5), 640–648. 10.1097/EDE.0b013e31818131e7

33. Jernigan, T. L., Brown, T. T., Hagler, D. J., Akshoomoff, N., Bartsch, H., Newman, E., Thompson, W. K., Bloss, C. S., Murray, S. S., Schork, N., Kennedy, D. N., Kuperman, J. M., McCabe, C., Chung, Y., Libiger, O., Maddox, M., Casey, B. J., Chang, L., Ernst, T. M., … Dale, A. M. (2016). The Pediatric Imaging, Neurocognition, and Genetics (PING) Data Repository. NeuroImage, 124, 1149– 1154. 10.1016/j.neuroimage.2015.04.057

34. Jørgensen, K. N., Nerland, S., Norbom, L. B., Doan, N. T., Nesvåg, R., Mørch-Johnsen, L., Haukvik, U. K., Melle, I., Andreassen, O. A., Westlye, L. T., & Agartz, I. (2016). Increased MRI-based cortical grey/white-matter contrast in sensory and motor regions in schizophrenia and bipolar disorder. Psychological Medicine, 46(9), 1971–1985. 10.1017/S0033291716000593

35. Khundrakpam, B., Choudhury, S., Vainik, U., AlLSharif, N., Bhutani, N., Jeon, S., Gold, I., & Evans, A. (2020). Distinct influence of parental occupation on cortical thickness and surface area in children and adolescents: Relation to selfLesteem. Human Brain Mapping, 41(18), 5097–5113. 10.1002/hbm.25169

36. Koenig, S. H. (1991). Cholesterol of myelin is the determinant of gray-white contrast in MRI of brain. Magnetic Resonance in Medicine, 20(2), 285–291. 10.1002/mrm.1910200210

37. Koenig, S. H., Brown III, R. D., Spiller, M., & Lundbom, N. (1990). Relaxometry of brain: Why white matter appears bright in MRI. Magnetic Resonance in Medicine, 14(3), 482–495. 10.1002/mrm.1910140306

38. Lebel, C., & Deoni, S. (2018). The development of brain white matter microstructure. NeuroImage, 182, 207–218. 10.1016/j.neuroimage.2017.12.097

39. Letourneau, N. L., Duffett-Leger, L., Levac, L., Watson, B., & Young-Morris, C. (2013). Socioeconomic Status and Child Development: A Meta-Analysis. Journal of Emotional and Behavioral Disorders, 21(3), 211–224. 10.1177/1063426611421007

40. Liu, S., Li, A., Zhu, M., Li, J., & Liu, B. (2019). Genetic influences on cortical myelination in the human brain. *Genes*, Brain and Behavior, 18(4), e12537. 10.1111/gbb.12537

41. Liu, W. M., Ali, S. R., Soleck, G., Hopps, J., dunston, K., & Pickett, T. (2004). Using Social Class in Counseling Psychology Research. Journal of Counseling Psychology, 51(1), 3–18. 10.1037/0022-0167.51.1.3

42. Long, K., & Renbarger, R. (2023). Persistence of Poverty: How Measures of Socioeconomic Status Have Changed Over Time. Educational Researcher, 0013189X2211414. 10.3102/0013189X221141409

43. Maxwell, S. E., & Cole, D. A. (2007). Bias in cross-sectional analyses of longitudinal mediation. Psychological Methods, 12(1), 23–44. 10.1037/1082-989X.12.1.23

44. Meyer, G. J., Finn, S. E., Eyde, L. D., Kay, G. G., Moreland, K. L., Dies, R. R., Eisman, E. J., Kubiszyn, T. W., & Reed, G. M. (2001). Psychological testing and psychological assessment: A review of evidence and issues. American Psychologist, 56(2), 128–165. 10.1037/0003-066X.56.2.128

45. Miller, K. L., Alfaro-Almagro, F., Bangerter, N. K., Thomas, D. L., Yacoub, E., Xu, J., Bartsch, A. J., Jbabdi, S., Sotiropoulos, S. N., Andersson, J. L. R., Griffanti, L., Douaud, G., Okell, T. W., Weale, P., Dragonu, I., Garratt, S., Hudson, S., Collins, R., Jenkinson, M., … Smith, S. M. (2016). Multimodal population brain imaging in the UK Biobank prospective epidemiological study. Nature Neuroscience, 19(11), 1523– 1536. 10.1038/nn.4393

46. Noble, K. G., Houston, S. M., Brito, N. H., Bartsch, H., Kan, E., Kuperman, J. M., Akshoomoff, N., Amaral, D. G., Bloss, C. S., Libiger, O., Schork, N. J., Murray, S. S., Casey, B. J., Chang, L., Ernst, T. M., Frazier, J. A., Gruen, J. R., Kennedy, D. N., Van Zijl, P., … Sowell, E. R. (2015). Family income, parental education and brain structure in children and adolescents. Nature Neuroscience, 18(5), 773–778. 10.1038/nn.3983

47. Norbom, L. B., Doan, N. T., Alnæs, D., Kaufmann, T., Moberget, T., Rokicki, J., Andreassen, O. A., Westlye, L. T., & Tamnes, C. K. (2019). Probing Brain Developmental Patterns of Myelination and Associations With Psychopathology in Youths Using Gray/White Matter Contrast. Biological Psychiatry, 85(5), 389–398. 10.1016/j.biopsych.2018.09.027

48. Norbom, L. B., Ferschmann, L., Parker, N., Agartz, I., Andreassen, O. A., Paus, T., Westlye, L. T., & Tamnes, C. K. (2021). New insights into the dynamic development of the cerebral cortex in childhood and adolescence: Integrating macro- and microstructural MRI findings. Progress in Neurobiology, 204, 102109. 10.1016/j.pneurobio.2021.102109

49. Norbom, L. B., Hanson, J., van der Meer, D., Ferschmann, L., Røysamb, E., von Soest, T., Andreassen, O. A., Agartz, I., Westlye, L. T., & Tamnes, C. K. (2022). Parental socioeconomic status is linked to cortical microstructure and language abilities in children and adolescents. Developmental Cognitive Neuroscience, 56, 101132. 10.1016/j.dcn.2022.101132

50. Norbom, L. B., Rokicki, J., Meer, D. van der, Alnæs, D., Doan, N. T., Moberget, T., Kaufmann, T., Andreassen, O. A., Westlye, L. T., & Tamnes, C. K. (2020). Testing relationships between multimodal modes of brain structural variation and age, sex and polygenic scores for neuroticism in children and adolescents. Translational Psychiatry, 10(1), 251. 10.1038/s41398-020-00931-1

51. Ørstavik, R. E., Czajkowski, N., Røysamb, E., Knudsen, G. P., Tambs, K., & Reichborn-Kjennerud, T. (2014). Sex Differences in Genetic and Environmental Influences on Educational Attainment and Income. Twin Research and Human Genetics, 17(6), 516–525. 10.1017/thg.2014.68

52. Petanjek, Z., Judas, M., Kostovic, I., & Uylings, H. B. M. (2008). Lifespan Alterations of Basal Dendritic Trees of Pyramidal Neurons in the Human Prefrontal Cortex: A Layer-Specific Pattern. Cerebral Cortex, 18(4), 915–929. 10.1093/cercor/bhm124

53. Petanjek, Z., Judaš, M., Šimić, G., Rašin, M. R., Uylings, H. B. M., Rakic, P., & Kostović, I. (2011). Extraordinary neoteny of synaptic spines in the human prefrontal cortex. Proceedings of the National Academy of Sciences, 108(32), 13281–13286. 10.1073/pnas.1105108108

54. Peter R. H. (1979). Synaptic density in human frontal cortex—Developmental changes and effects of aging. Brain Research, 163(2), 195–205. 10.1016/0006-8993(79)90349-4

55. Piccolo, L. R., Merz, E. C., He, X., Sowell, E. R., Noble, K. G., & Pediatric Imaging, Neurocognition, Genetics Study. (2016). Age-Related Differences in Cortical Thickness Vary by Socioeconomic Status. PLOS ONE, 11(9), e0162511. 10.1371/journal.pone.0162511

56. Rakesh, D., & Whittle, S. (2021). Socioeconomic status and the developing brain – A systematic review of neuroimaging findings in youth. Neuroscience & Biobehavioral Reviews, 130, 379–407. 10.1016/j.neubiorev.2021.08.027

57. Rakesh, D., Zalesky, A., & Whittle, S. (2022). Assessment of Parent Income and Education, Neighborhood Disadvantage, and Child Brain Structure. JAMA Network Open, 5(8), e2226208. 10.1001/jamanetworkopen.2022.26208

58. Rakic, P. (1988). Specification of cerebral cortical areas. Science, 241(4862), 170.

59. Reiss, F. (2013). Socioeconomic inequalities and mental health problems in children and adolescents: A systematic review. Social Science & Medicine, 90, 24–31. 10.1016/j.socscimed.2013.04.026

60. Rosen, A. F. G., Roalf, D. R., Ruparel, K., Blake, J., Seelaus, K., Villa, L. P., Ciric, R., Cook, P. A., Davatzikos, C., Elliott, M. A., Garcia de La Garza, A., Gennatas, E. D., Quarmley, M., Schmitt, J. E., Shinohara, R. T., Tisdall, M. D., Craddock, R. C., Gur, R. E., Gur, R. C., & Satterthwaite, T. D. (2018). Quantitative assessment of structural image quality. NeuroImage, 169, 407–418. 10.1016/j.neuroimage.2017.12.059

61. Salat, D. H., Lee, S. Y., van der Kouwe, A. J., Greve, D. N., Fischl, B., & Rosas, H. D. (2009). Age-associated alterations in cortical gray and white matter signal intensity and gray to white matter contrast. NeuroImage, 48(1), 21–28. PubMed. 10.1016/j.neuroimage.2009.06.074

62. Seldon, H. L. (2005). Does brain white matter growth expand the cortex like a balloon? Hypothesis and consequences. *Laterality: Asymmetries of Body*, Brain and Cognition, 10(1), 81–95. 10.1080/13576500342000310

63. Sirin, S. R. (2005). Socioeconomic Status and Academic Achievement: A Meta-Analytic Review of Research. Review of Educational Research, 75(3), 417–453. 10.3102/00346543075003417

64. Strike, L. T., Hansell, N. K., Couvy-Duchesne, B., Thompson, P. M., de Zubicaray, G. I., McMahon, K. L., & Wright, M. J. (2019). Genetic Complexity of Cortical Structure: Differences in Genetic and Environmental Factors Influencing Cortical Surface Area and Thickness. Cerebral Cortex, 29(3), 952–962. 10.1093/cercor/bhy002

65. Sydnor, V. J., Larsen, B., Bassett, D. S., Alexander-Bloch, A., Fair, D. A., Liston, C., Mackey, A. P., Milham, M. P., Pines, A., Roalf, D. R., Seidlitz, J., Xu, T., Raznahan, A., & Satterthwaite, T. D. (2021). Neurodevelopment of the association cortices: Patterns, mechanisms, and implications for psychopathology. Neuron, 109(18), 2820– 2846. 10.1016/j.neuron.2021.06.016

66. Tambs, K., Kendler, K. S., Reichborn-Kjennerud, T., Aggen, S. H., Harris, J. R., Neale, M. C., Hettema, J. M., Sundet, J. M., Battaglia, M., & Røysamb, E. (2012). Genetic and environmental contributions to the relationship between education and anxiety disorders - a twin study: Common genes for education and anxiety. Acta Psychiatrica Scandinavica, 125(3), 203–212. 10.1111/j.1600-0447.2011.01799.x

67. Taylor, R. L., Cooper, S. R., Jackson, J. J., & Barch, D. M. (2020). Assessment of Neighborhood Poverty, Cognitive Function, and Prefrontal and Hippocampal Volumes in Children. JAMA Network Open, 3(11), e2023774. 10.1001/jamanetworkopen.2020.23774

68. Thomas, M. S. C., & Coecke, S. (2023). Associations between Socioeconomic Status, Cognition, and Brain Structure: Evaluating Potential Causal Pathways Through Mechanistic Models of Development. Cognitive Science, 47(1). 10.1111/cogs.13217

69. Tomasi, D., & Volkow, N. D. (2021). Associations of family income with cognition and brain structure in USA children: Prevention implications. Molecular Psychiatry, 26(11), 6619–6629. 10.1038/s41380-021-01130-0

70. Vargas, T., Damme, K. S. F., & Mittal, V. A. (2020). Neighborhood deprivation, prefrontal morphology and neurocognition in late childhood to early adolescence. NeuroImage, 220, 117086. 10.1016/j.neuroimage.2020.117086

71. Vidal-Pineiro, D., Parker, N., Shin, J., French, L., Grydeland, H., Jackowski, A. P., Mowinckel, A. M., Patel, Y., Pausova, Z., Salum, G., Sørensen, Ø., Walhovd, K. B., Paus, T., Fjell, A. M., & the Alzheimer’s Disease Neuroimaging Initiative and the Australian Imaging Biomarkers and Lifestyle flagship study of ageing. (2020). Cellular correlates of cortical thinning throughout the lifespan. Scientific Reports, 10(1), 21803. 10.1038/s41598-020-78471-3

72. von Stumm, S., & Plomin, R. (2015). Socioeconomic status and the growth of intelligence from infancy through adolescence. Intelligence, 48, 30–36. 10.1016/j.intell.2014.10.002

73. Waxman, S. G., & Bennett, M. V. L. (1972). Relative Conduction Velocities of Small Myelinated and Non-myelinated Fibres in the Central Nervous System. Nature New Biology, 238(85), 217–219. 10.1038/newbio238217a0

74. Wolfers, T., Arenas, A. L., Onnink, A. M. H., Dammers, J., Hoogman, M., Zwiers, M. P., Buitelaar, J. K., Franke, B., Marquand, A. F., & Beckmann, C. F. (2017). Refinement by integration: Aggregated effects of multimodal imaging markers on adult ADHD. Journal of Psychiatry & Neuroscience, 42(6), 386–394. 10.1503/jpn.160240

75. Yosso, T. (2005). Whose cultural has capital. A Critical Race Theory Discussion Of.

76. Zhang, H., Lee, Z. X., White, T., & Qiu, A. (2020). Parental and social factors in relation to child psychopathology, behavior, and cognitive function. Translational Psychiatry, 10(1), 80. 10.1038/s41398-020-0761-6

