## Supplementary Information for "Parental education and income are linked to offspring cortical brain structure and psychopathology at 9-11 years"

**Supplemental Information (SI)**

*Participants*

All ABCD data is stored in the NIMH Data Archive Collection #2573, which is available for registered and authorised users (Request #7474, PI: Westlye). The 4.0 release will be permanently available as a persistent dataset defined in the NDA Study 1299 and has been assigned the DOI 10.15154/1523041.

Participants were generally recruited through the school system, and was informed by sex, ethnicity, SES and urbanicity, in an attempt to address selection biases. Parental informed consent as well as child assent was obtained for all subjects.

ABCD Study^®^ exclusion criteria included non-English proficiency, contraindications for MRI, a history of major neurological disorders, extremely premature birth, and a diagnosis of schizophrenia, substance abuse disorder or moderate to severe autism spectrum disorder (Karcher et al., 2018). The Institutional Review Board at the University of California, San Diego, approved all aspects of ABCD Study^®^ (Auchter et al., 2018)^.^ The current study was conducted in line with the Declaration of Helsinki and was approved by the Regional Committee for Medical and Health Research Ethics (REK 2019/943).

*Measurements of socioeconomic status*

A partner was defined as “any significant figure that helps you in raising your child or has helped you for more than 2 years and is =>40% involved in the child’s daily activities, i.e., a spouse, boyfriend/girlfriend or relative”.

Parental education was assessed by ranking the question “What is the highest grade or level of school you have completed, or the highest degree you have received” on a 22-point scale. The scale was recoded to years of total education resulting in recoding of levels 13-19, as presented in SI Table 1.

Parental income was assessed by scoring the question “How much did you earn, before taxes and other deductions, during the past 12 months” on a 10-point scale. Total combined income was assessed by scoring the question “What is your total combined family income for the past 12 months including all sources?” on the same 10-point scale. The scale was recoded as the median point of each bracket as presented in SI Table 1. The maximum of the last bracket was pragmatically set to 350.000$ based on online searches reporting that the threshold for a US household to be within the top 1% in 2018 was 434,454.80$ (<https://dqydj.com/2018-average-median-top-household-income-percentiles/>).

Our main objective was to assess the relations between multimodally fused cortical macro- and microstructure and conventional SES subfactors. While we had access to occupational information on a 22-point scale, the scaling was not based on any form of status-based ranking. To maintain the integrity of our SES measures and avoid introducing subjective bias, we chose to not include assessments of occupation.

| Question | Representation of numbers | Recoding |
| --- | --- | --- |
| What is the highest grade or level of school you have completed or the highest degree you have received? | 0= Never attended/Kindergarten only  1= 1st grade  2= 2nd grade  3= 3rd grade  4= 4th grade  5= 5th grade  6= 6th grade  7= 7th grade  8= 8th grade 8  9= 9th grade  10= 10th grade  11= 11th grade  12= 12th grade  13= High school graduate  14= GED or equivalent  15= Some college  16= Associate degree: Occupational  17= Associate degree: Academic Program  18= Bachelor's degree (i.e. BA)  19= Master's degree (i.e. MA)  20= Professional School degree (i.e. MD)  21= Doctoral degree (i.e. PhD) | 0= 0 years of education  1= 1 year of education  2= 2 years of education  3= 3 years of education  4= 4 years of education  5= 5 years of education  6= 6 years of education  7= 7 years of education  8= 8 years of education  9= 9 years of education  10= 10 years of education  11= 11 years of education  12= 12 years of education  13= 12 years of education  14= 12 years of education  15= 14 years of education  16= 14 years of education  17= 14 years of education  18= 16 years of education  19= 18 years of education  20= 20 years of education  21= 21 years of education |
| How much did you earn, before taxes and other deductions, during the past 12 months? | 1= Less than $5,000  2= $5,000 through $11,999  3= $12,000 through $15,999  4= $16,000 through $24,999  5= $25,000 through $34,999  6= $35,000 through $49,999  7= $50,000 through $74,999  8= $75,000 through $99,999  9= $100,000 through $199,999;  10= $200,000 and greater  777= Refuse to answer  999= Don't know | 1= $2500  2= $8499,5  3= $13999,5  4= $20499,5  5= $29999,5  6= $42499,5  7= $62499,5  8= $87499,5  9= $149999,5  10=275000  777= NA  999= NA |
| What is your total combined family income for the past 12 months? This should include income from all sources, wages, rent from properties, social security, disability and/or veteran's benefits, unemployment benefits, help from relative and so on | 1= Less than $5,000  2= $5,000 through $11,999  3= $12,000 through $15,999  4= $16,000 through $24,999  5= $25,000 through $34,999  6= $35,000 through $49,999  7= $50,000 through $74,999  8= $75,000 through $99,999  9= $100,000 through $199,999;  10= $200,000 and greater  777= Refuse to answer  999= Don't know | 1= $2500  2= $8499,5  3= $13999,5  4= $20499,5  5= $29999,5  6= $42499,5  7= $62499,5  8= $87499,5  9= $149999,5  10=275000  777= NA  999= NA |

SI Table 1. Parental education and parental income details. The table shows the socioeconomic status related questions presented to the parents, its numeric levels, and what these levels represent.

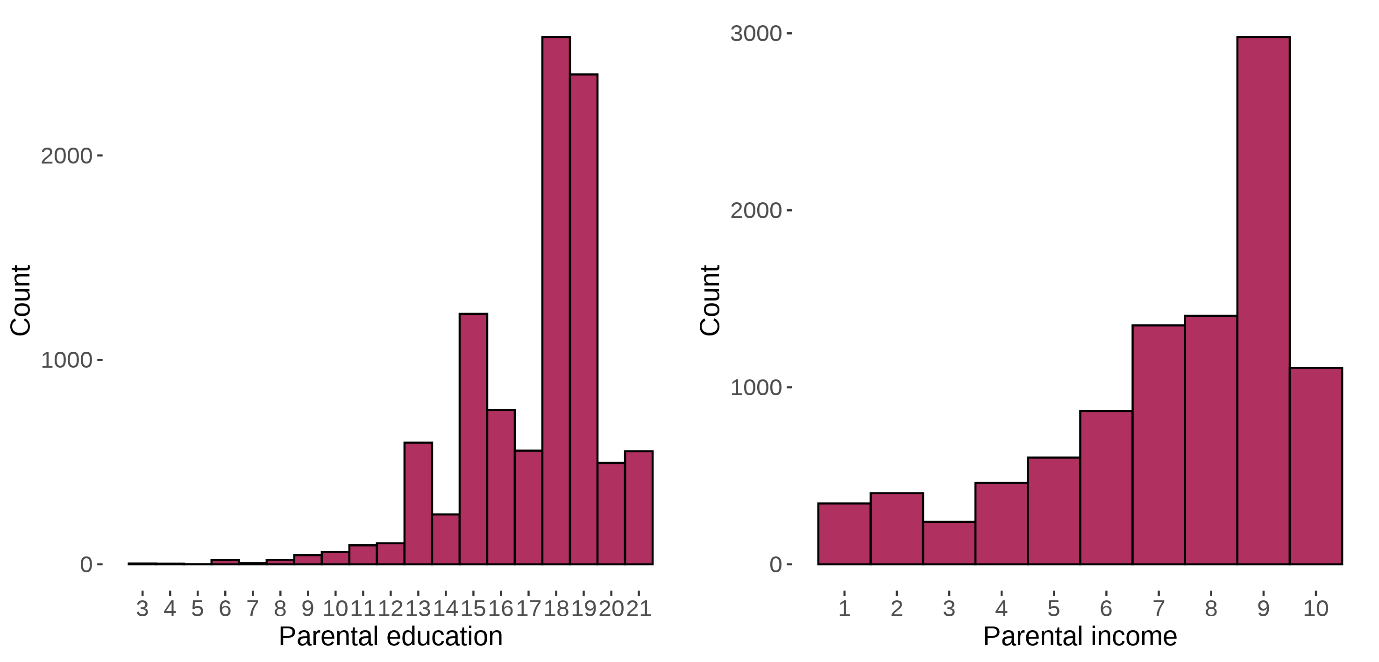

SI Figure 1. Parental SES variables. The figure depicts the distributions of raw parental education and parental income scores for the final sample (n=9758).

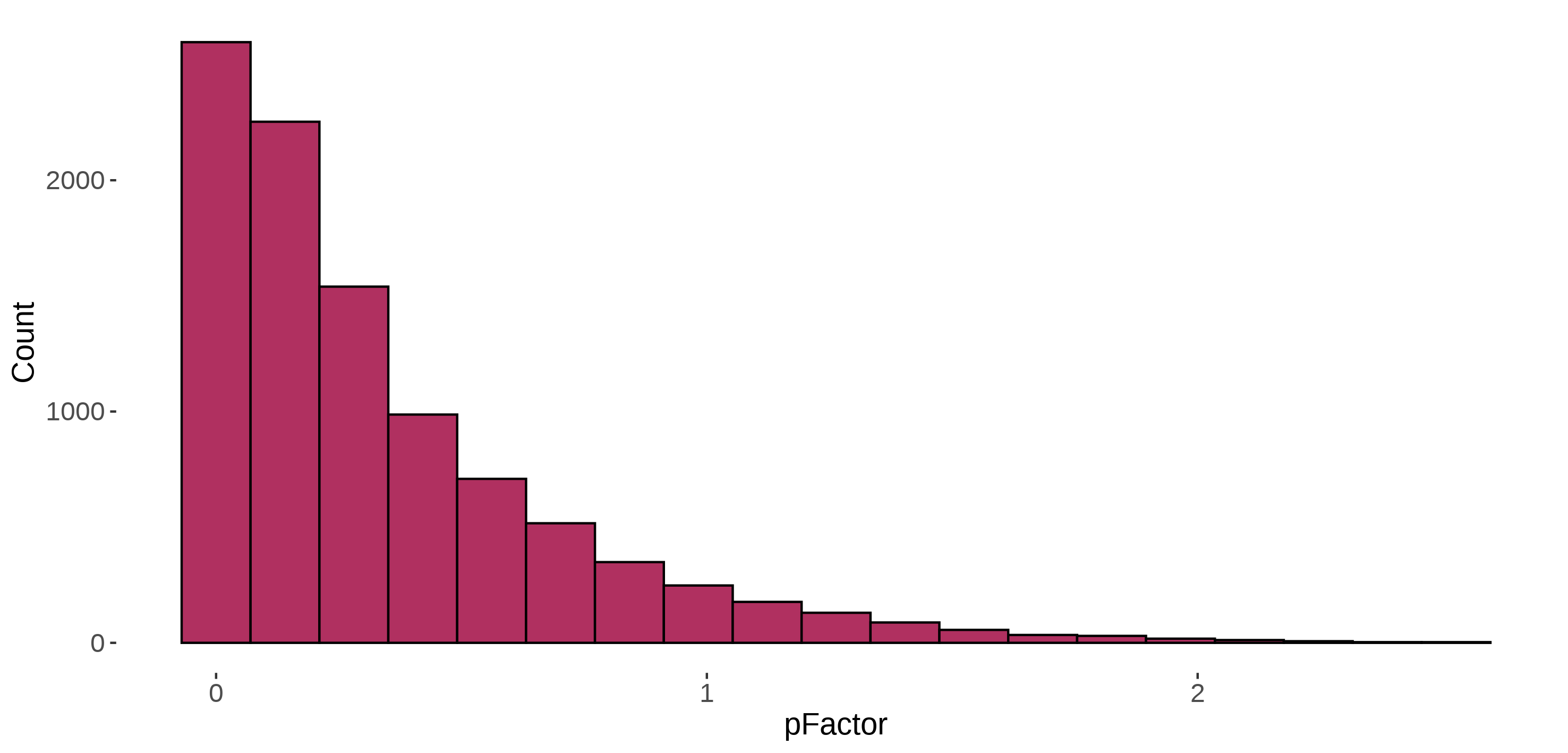

SI Figure 2. Child psychopathology. The figure depicts the distribution of child p-factor scores within the final sample (n=9758).

*FreeSurfer based macro-and microstructural surface calculations*

Cortical thickness was computed as the shortest vertex-wise distance between the white and pial surface, while vertex-wise surface area was calculated by summing the triangle areas converging at each particular vertex on the white surface. Cortical curvature based on the white surface, was calculated as the vertex wise average of the two principal curvature directions, so that higher curvature represents a decrease in curvature radius, usually indicating sharper folding. GWC was calculated using intensity sampling from “rawavg.mgz”, where white matter was sampled 1 mm below-, and grey matter was sampled 30% above the white surface. The vertex wise percentage difference was then computed as 100 × (white − grey)/[(white + grey)/2], so that lower GWC reflects more similar grey and white matter, a pattern documented across youth development.

*Scanner harmonization*

Smoothed cortical surfaces were imported to R and the package neuroCombat (Fortin et al., 2018) was employed at vertex level to harmonize data across 29 scanners. We included age, sex, p-factor, parental education, and parental income as co-variates to our model to preserve variance related to these variables during the harmonization procedure. The spread of global cortical measures across scanners, pre- and post-neuroCombat adjustment is presented in SI Figures 3-6.

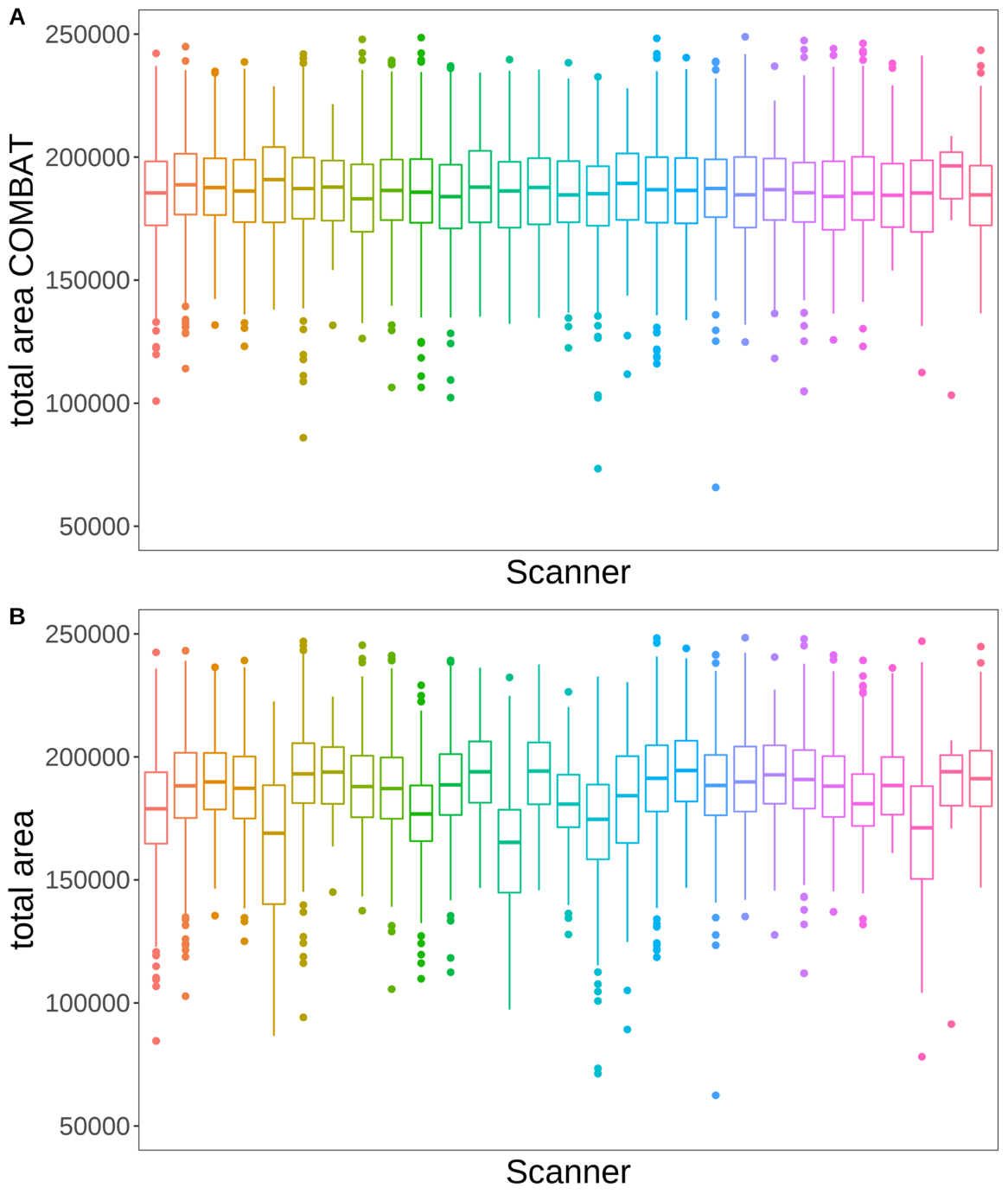

SI Figure 3. Box plots of total surface area across scanners pre- and post harmonization. The figure shows box plots of the distribution of total surface area across scanners post scanner harmonization at the top (A), and pre harmonization at the bottom (B).

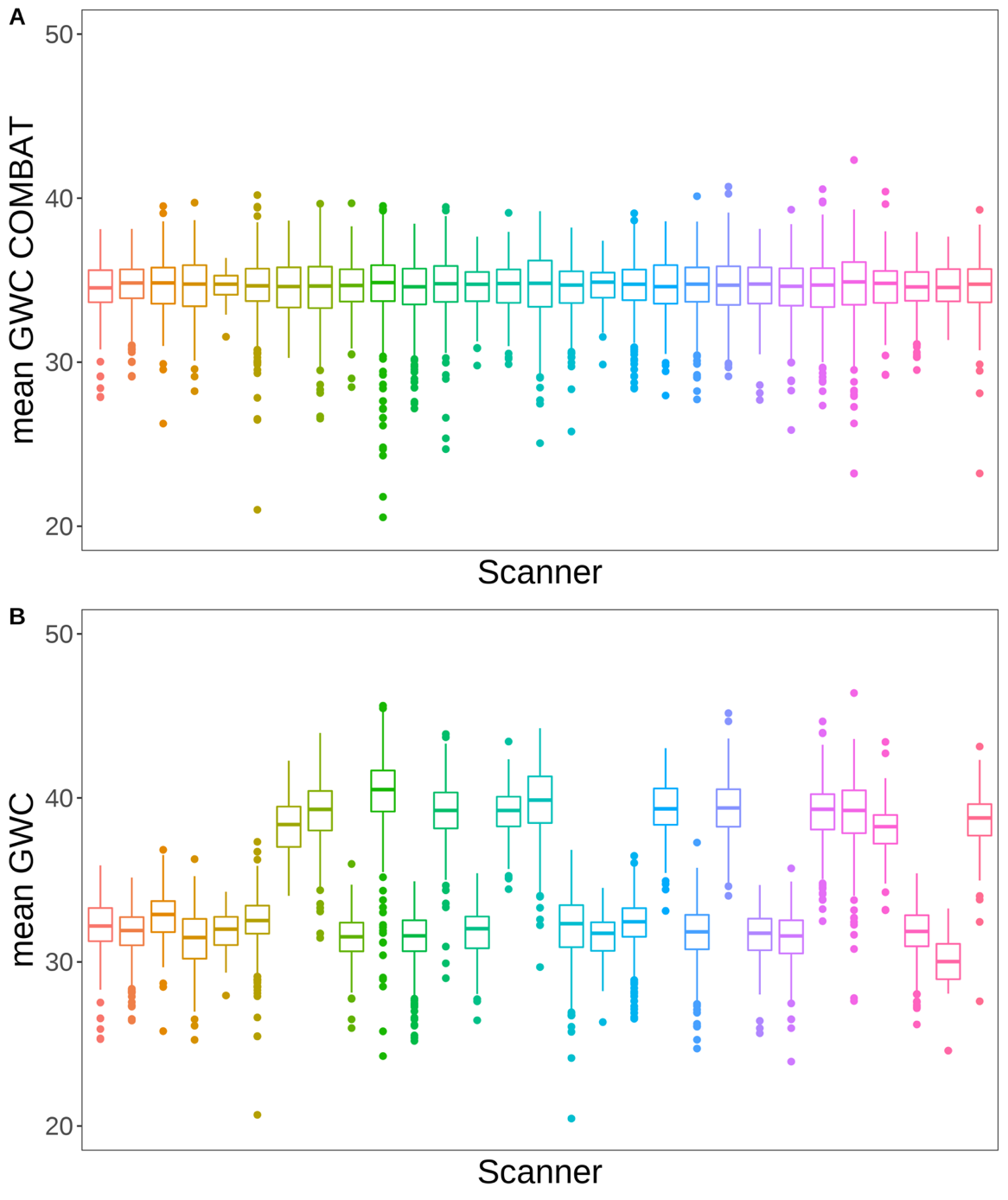

SI Figure 4. Box plots of mean grey/white-matter contrast (GWC) across scanners pre- and post harmonization. The figure shows box plots of the distribution of mean GWC across scanners post scanner harmonization at the top (A), and pre harmonization at the bottom (B).

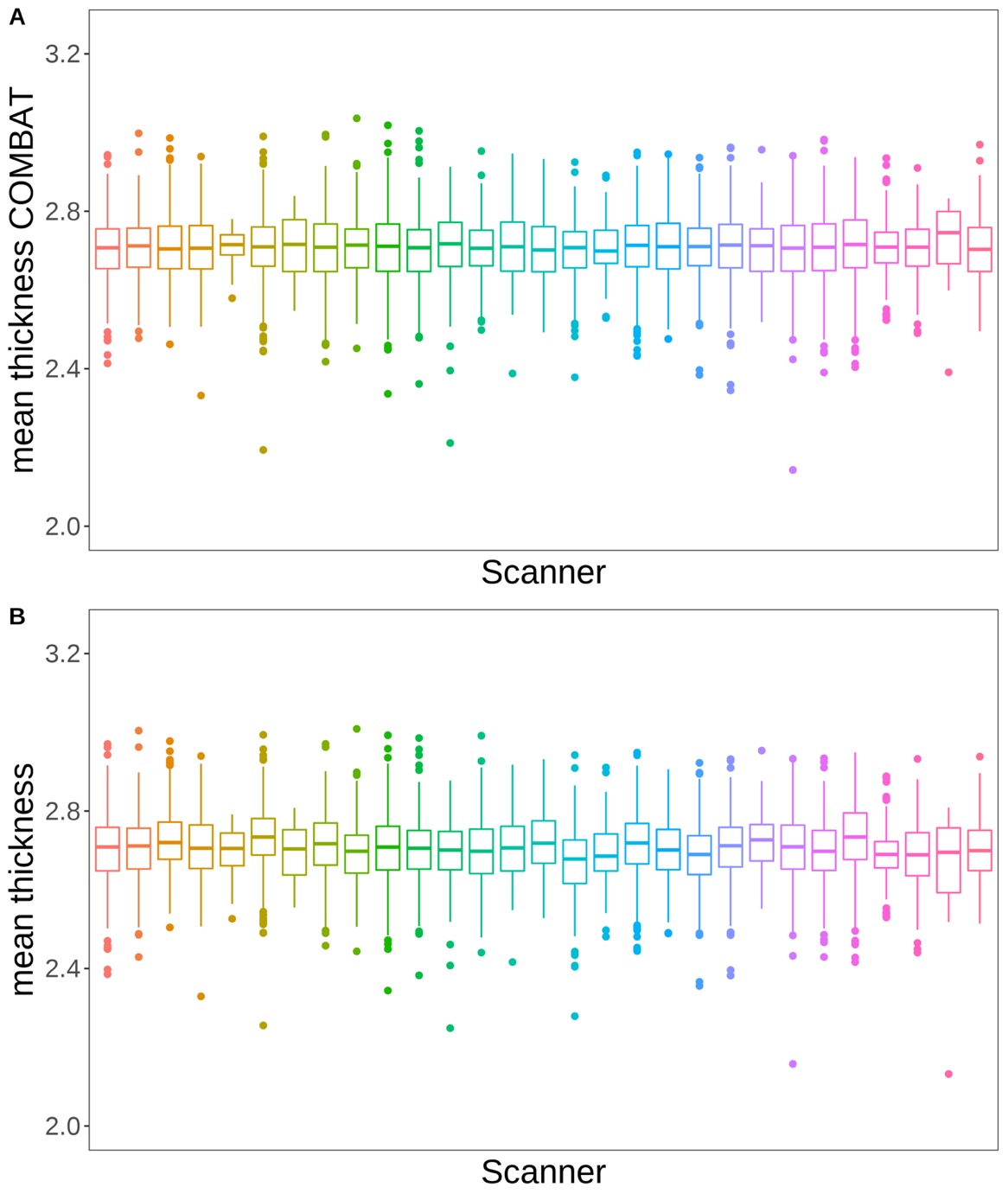

SI Figure 5. Box plots of mean cortical thickness across scanners pre- and post harmonization. The figure shows box plots of the distribution of mean cortical thickness across scanners post scanner harmonization at the top (A), and pre harmonization at the bottom (B).

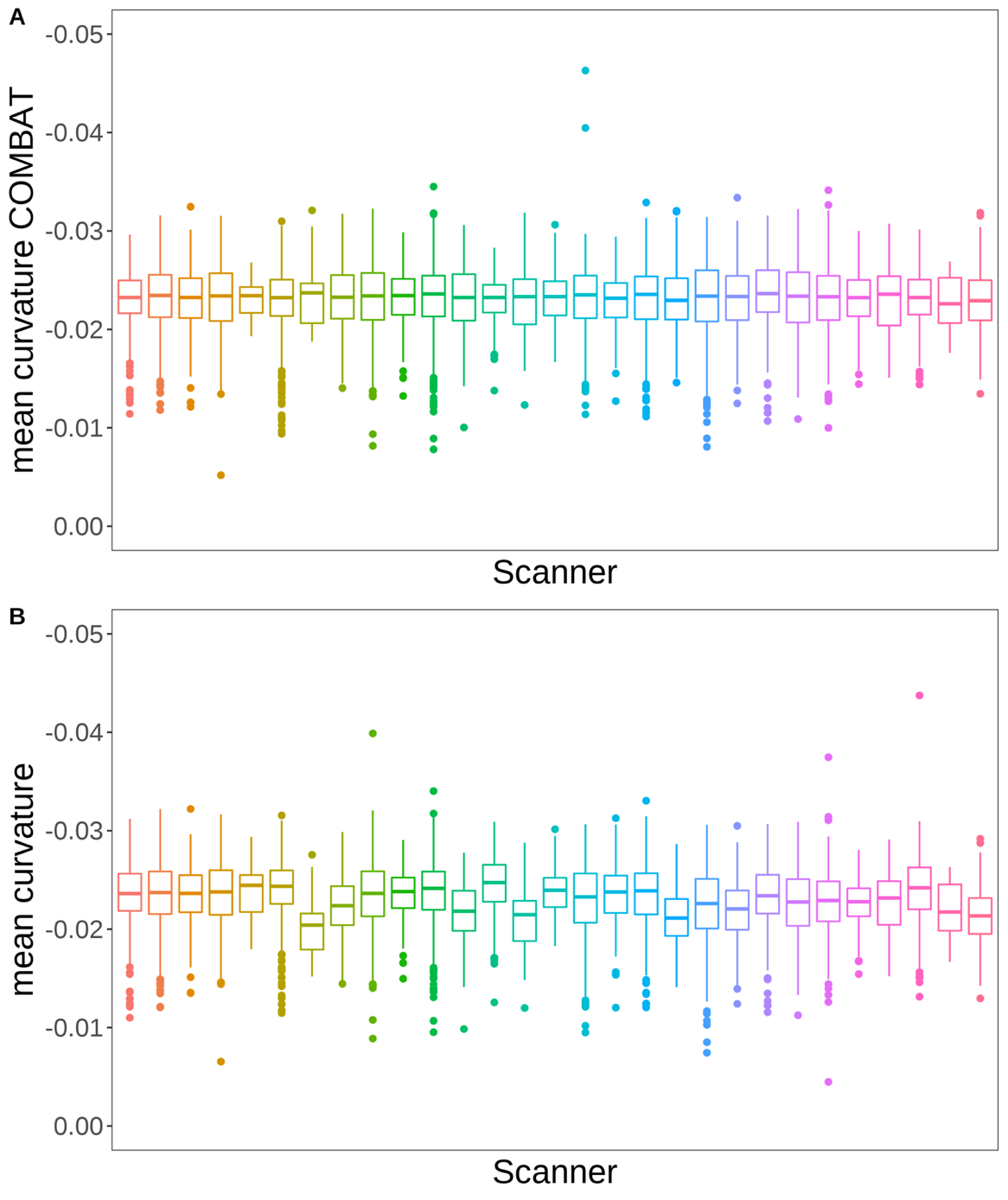

SI Figure 6. Box plots of mean curvature across scanners pre- and post harmonization. The figure shows box plots of the distribution of mean curvature across scanners post scanner harmonization at the top (A), and pre harmonization at the bottom (B).

*Statistical analyses*

We tested the quadratic relationship between parental education and each of the 70 components, separately. The second-degree polynomial of parental education was included as an independent variable while linked independent component analysis (LICA) subject loading was included as a dependent variable. The first-degree polynomial of parental education age, sex, and the 4 genetic ancestry factors (GAFs) were added as fixed effects, while family ID and monozygotic twin status were added as random effects. We then ran identical analyses testing the quadratic relationship between parental income and each LICA subject loading.

**Results**

*Nonlinear associations between parental socioeconomic status and youth cortical structure*

Linear mixed effects (LME) models with SES related polynomials of degree 2, revealed no quadratic relationships between parental education and youth cortical structure. Parental income showed a negative and positive quadratic relationship with IC33 and IC54, respectively.

There was a significant negative quadratic relationship between parental income and IC33, with no significant main effects being reported between the two. IC33 was dominated by a bi-hemispheric regional surface area (76%) pattern, specifically of larger insular area, extending into the temporal and parietal and frontal lobes, as well as smaller medial area extending into lateral occipital-, parietal and frontal lobes. IC33 also showed bidirectional and bi-hemispheric local patterns adjacent to the insula for thickness (9%), GWC (8%), and curvature (7%) (Figure 3). Visualizations of the inverted U-association as presented in SI Figure 7, indicate a positive association between parental income and local multimodal insular patterns in the low end of the income scale, a pattern that reverses in the higher end of the scale.

There was a significant positive quadratic relationship between parental income and IC54, and analyses also showed a significant negative main effect between the two. As presented in the main text IC54 had a general bi-hemispheric pattern of larger occipital and smaller frontal surface area (77%), larger medial occipital thickness (13%), and higher medial occipital, and lower lateral occipital GWC (10%) (Figure 3). Visualizations of the U-association as presented in SI Figure 7, indicates an exponential decay in the association between parental income and local variations in child surface area, GWC, and thickness. Our results indicate that after certain levels of monetary security, additional increases in income show weaker associations with child cortical structure.

|  | PARENTAL EDUCATION | | | | | PARENTAL INCOME | | | |
| --- | --- | --- | --- | --- | --- | --- | --- | --- | --- |
| IC | Cohens D | T statistic | | p-value (uncorrected) | p-value (corrected) | Cohens D | T statistic | p-value (uncorrected) | p-value (corrected) |
| 1 | 0 | | 0.11 | 0.911 | 0.994 | -0.04 | -1.7 | 0.089 | 0.986 |
| 2 | 0 | | 0.23 | 0.817 | 0.994 | -0.03 | -1.29 | 0.197 | 0.986 |
| 3 | 0.02 | | 0.86 | 0.391 | 0.994 | -0.04 | -1.63 | 0.103 | 0.986 |
| 4 | -0.02 | | -0.95 | 0.34 | 0.994 | 0 | 0.21 | 0.835 | 0.986 |
| 5 | -0.02 | | -0.83 | 0.409 | 0.994 | -0.04 | -1.78 | 0.076 | 0.986 |
| 6 | -0.01 | | -0.41 | 0.68 | 0.994 | 0.02 | 0.78 | 0.433 | 0.986 |
| 7 | 0 | | 0.21 | 0.836 | 0.994 | 0.02 | 0.91 | 0.365 | 0.986 |
| 8 | 0.02 | | 0.84 | 0.401 | 0.994 | 0.02 | 0.8 | 0.424 | 0.986 |
| 9 | 0.01 | | 0.36 | 0.718 | 0.994 | 0.02 | 0.97 | 0.331 | 0.986 |
| 10 | 0.03 | | 1.59 | 0.111 | 0.994 | -0.01 | -0.49 | 0.623 | 0.986 |
| 11 | 0.04 | | 1.78 | 0.075 | 0.994 | -0.03 | -1.26 | 0.208 | 0.986 |
| 12 | 0 | | -0.03 | 0.974 | 0.994 | 0 | 0.02 | 0.986 | 0.986 |
| 13 | 0 | | -0.04 | 0.971 | 0.994 | 0.01 | 0.27 | 0.786 | 0.986 |
| 14 | -0.01 | | -0.6 | 0.55 | 0.994 | 0 | 0.18 | 0.856 | 0.986 |
| 15 | -0.01 | | -0.57 | 0.571 | 0.994 | -0.01 | -0.33 | 0.739 | 0.986 |
| 16 | 0.02 | | 0.86 | 0.389 | 0.994 | 0.02 | 0.93 | 0.351 | 0.986 |
| 17 | -0.01 | | -0.6 | 0.549 | 0.994 | -0.03 | -1.41 | 0.158 | 0.986 |
| 18 | 0.01 | | 0.56 | 0.577 | 0.994 | 0.05 | 2.34 | 0.019* | 0.986 |
| 19 | 0.03 | | 1.17 | 0.24 | 0.994 | -0.01 | -0.31 | 0.754 | 0.986 |
| 20 | 0.04 | | 1.69 | 0.09 | 0.994 | 0 | -0.09 | 0.93 | 0.986 |
| 21 | -0.01 | | -0.41 | 0.68 | 0.994 | -0.03 | -1.49 | 0.136 | 0.986 |
| 22 | -0.01 | | -0.37 | 0.709 | 0.994 | -0.02 | -0.94 | 0.349 | 0.986 |
| 23 | 0.03 | | 1.47 | 0.142 | 0.994 | 0.01 | 0.41 | 0.682 | 0.986 |
| 24 | 0.02 | | 0.78 | 0.435 | 0.994 | 0.02 | 1.08 | 0.281 | 0.986 |
| 25 | -0.01 | | -0.3 | 0.761 | 0.994 | 0.02 | 1.07 | 0.284 | 0.986 |
| 26 | -0.01 | | -0.28 | 0.779 | 0.994 | -0.01 | -0.35 | 0.725 | 0.986 |
| 27 | -0.03 | | -1.32 | 0.188 | 0.994 | 0.02 | 0.81 | 0.419 | 0.986 |
| 28 | 0 | | -0.2 | 0.839 | 0.994 | -0.04 | -1.61 | 0.107 | 0.986 |
| 29 | -0.01 | | -0.65 | 0.518 | 0.994 | 0.03 | 1.52 | 0.129 | 0.986 |
| 30 | -0.05 | | -2.43 | 0.015* | 0.994 | -0.02 | -0.8 | 0.423 | 0.986 |
| 31 | -0.01 | | -0.35 | 0.727 | 0.994 | -0.01 | -0.4 | 0.689 | 0.986 |
| 32 | 0.02 | | 0.87 | 0.386 | 0.994 | -0.02 | -1 | 0.316 | 0.986 |
| 33 | -0.05 | | -2.45 | 0.014* | 0.961 | -0.08 | -3.64 | <0.000* | 0.019* |
| 34 | 0.02 | | 1.06 | 0.291 | 0.994 | 0 | -0.13 | 0.9 | 0.986 |
| 35 | 0.04 | | 1.85 | 0.065 | 0.994 | 0 | 0.23 | 0.822 | 0.986 |
| 36 | 0.07 | | 3.29 | 0.001* | 0.071 | 0.03 | 1.24 | 0.214 | 0.986 |
| 37 | -0.01 | | -0.45 | 0.652 | 0.994 | 0.04 | 1.81 | 0.07 | 0.986 |
| 38 | -0.02 | | -1.02 | 0.31 | 0.994 | 0.01 | 0.36 | 0.717 | 0.986 |
| 39 | 0.04 | | 2.08 | 0.037* | 0.994 | 0.04 | 1.64 | 0.1 | 0.986 |
| 40 | -0.02 | | -1.1 | 0.272 | 0.994 | 0.01 | 0.34 | 0.736 | 0.986 |
| 41 | 0 | | 0.15 | 0.88 | 0.994 | 0.01 | 0.46 | 0.646 | 0.986 |
| 42 | 0.01 | | 0.6 | 0.551 | 0.994 | 0.02 | 1.05 | 0.293 | 0.986 |
| 43 | 0 | | 0.13 | 0.896 | 0.994 | 0.05 | 2.08 | 0.038* | 0.986 |
| 44 | 0.01 | | 0.7 | 0.487 | 0.994 | 0.05 | 2.24 | 0.025* | 0.986 |
| 45 | 0.02 | | 0.84 | 0.404 | 0.994 | -0.03 | -1.47 | 0.143 | 0.986 |
| 46 | 0.04 | | 1.86 | 0.064 | 0.994 | 0.02 | 1.07 | 0.282 | 0.986 |
| 47 | 0.01 | | 0.59 | 0.554 | 0.994 | 0.04 | 1.98 | 0.048* | 0.986 |
| 48 | 0.01 | | 0.25 | 0.802 | 0.994 | -0.04 | -1.85 | 0.065 | 0.986 |
| 49 | -0.01 | | -0.69 | 0.492 | 0.994 | 0.02 | 0.81 | 0.419 | 0.986 |
| 50 | 0.01 | | 0.36 | 0.718 | 0.994 | -0.01 | -0.51 | 0.612 | 0.986 |
| 51 | 0 | | -0.01 | 0.994 | 0.994 | 0 | -0.17 | 0.864 | 0.986 |
| 52 | 0 | | -0.15 | 0.885 | 0.994 | 0 | 0.19 | 0.846 | 0.986 |
| 53 | 0.02 | | 0.89 | 0.375 | 0.994 | -0.02 | -0.85 | 0.396 | 0.986 |
| 54 | 0.04 | | 1.79 | 0.073 | 0.994 | 0.08 | 3.53 | <0.000* | 0.029* |
| 55 | 0.02 | | 1.14 | 0.254 | 0.994 | 0 | -0.04 | 0.966 | 0.986 |
| 56 | 0.02 | | 1.16 | 0.245 | 0.994 | -0.01 | -0.65 | 0.515 | 0.986 |
| 57 | -0.02 | | -0.78 | 0.436 | 0.994 | -0.01 | -0.57 | 0.572 | 0.986 |
| 58 | 0.02 | | 1.01 | 0.31 | 0.994 | 0 | 0.05 | 0.962 | 0.986 |
| 59 | 0.08 | | - | - | - | -0.1 | - | - | - |
| 60 | 0.04 | | 1.74 | 0.082 | 0.994 | -0.04 | -1.87 | 0.061 | 0.986 |
| 61 | 0.02 | | 1.03 | 0.301 | 0.994 | 0.06 | 2.55 | 0.011* | 0.725 |
| 62 | -0.04 | | -1.74 | 0.083 | 0.994 | -0.01 | -0.51 | 0.608 | 0.986 |
| 63 | -0.03 | | -1.33 | 0.182 | 0.994 | -0.02 | -0.95 | 0.342 | 0.986 |
| 64 | 0 | | -0.02 | 0.981 | 0.994 | -0.01 | -0.65 | 0.517 | 0.986 |
| 65 | 0 | | -0.11 | 0.91 | 0.994 | -0.01 | -0.29 | 0.768 | 0.986 |
| 66 | 0.01 | | 0.66 | 0.51 | 0.994 | -0.01 | -0.41 | 0.683 | 0.986 |
| 67 | -0.03 | | -1.36 | 0.173 | 0.994 | -0.01 | -0.28 | 0.776 | 0.986 |
| 68 | 0.03 | | 1.44 | 0.15 | 0.994 | -0.01 | -0.37 | 0.71 | 0.986 |
| 69 | -0.03 | | -1.22 | 0.224 | 0.994 | -0.03 | -1.37 | 0.171 | 0.986 |
| 70 | -0.02 | | -0.74 | 0.46 | 0.994 | -0.03 | -1.37 | 0.172 | 0.986 |

SI Table 2. Associations between parental socioeconomic status (SES) polynomial of degree 2 and component loadings. The table depicts Cohens D, T statistic, and uncorrected- and FDR-corrected p-values from the statistical analyses of the associations between parental education, parental income, and each component loading. Significant p-values are marked with Asterix (*).

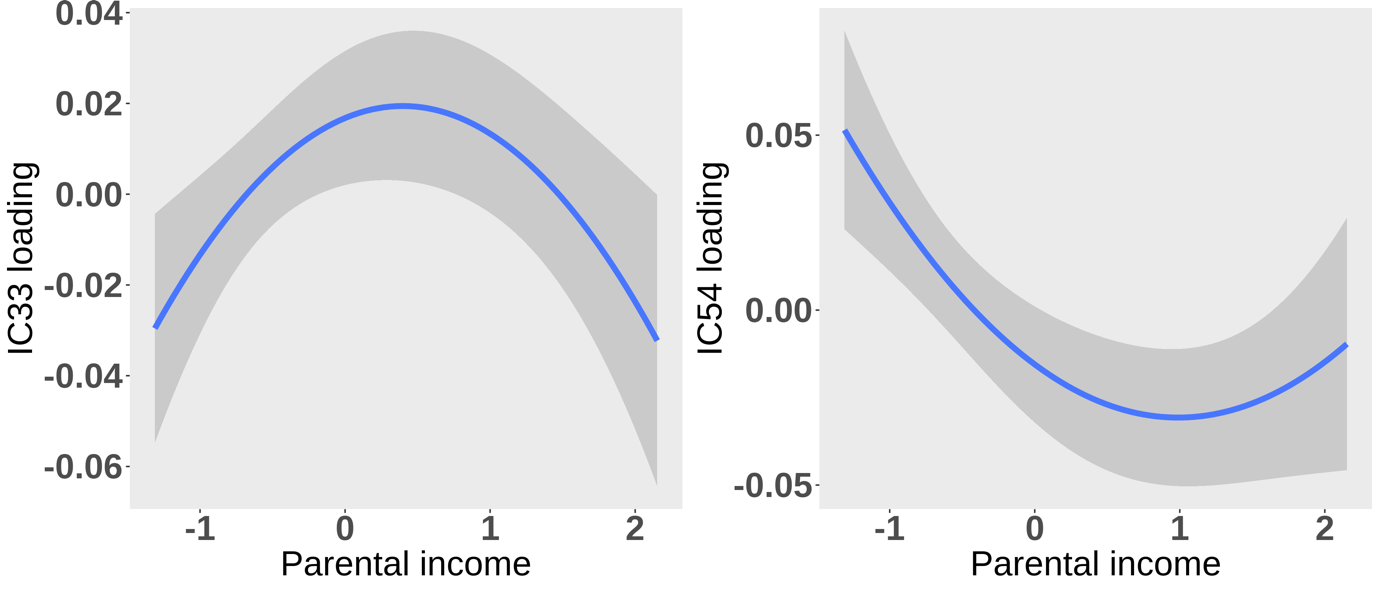

SI Figure 7. Visualization of the nonlinear relationship between parental income and IC33 and IC54. The figure is a visualization of the quadratic relationship between parental income and IC33 (left) and IC54 (right). IC loadings are residualized by all co-variates from our statistical model, apart from the main effect of each IC.

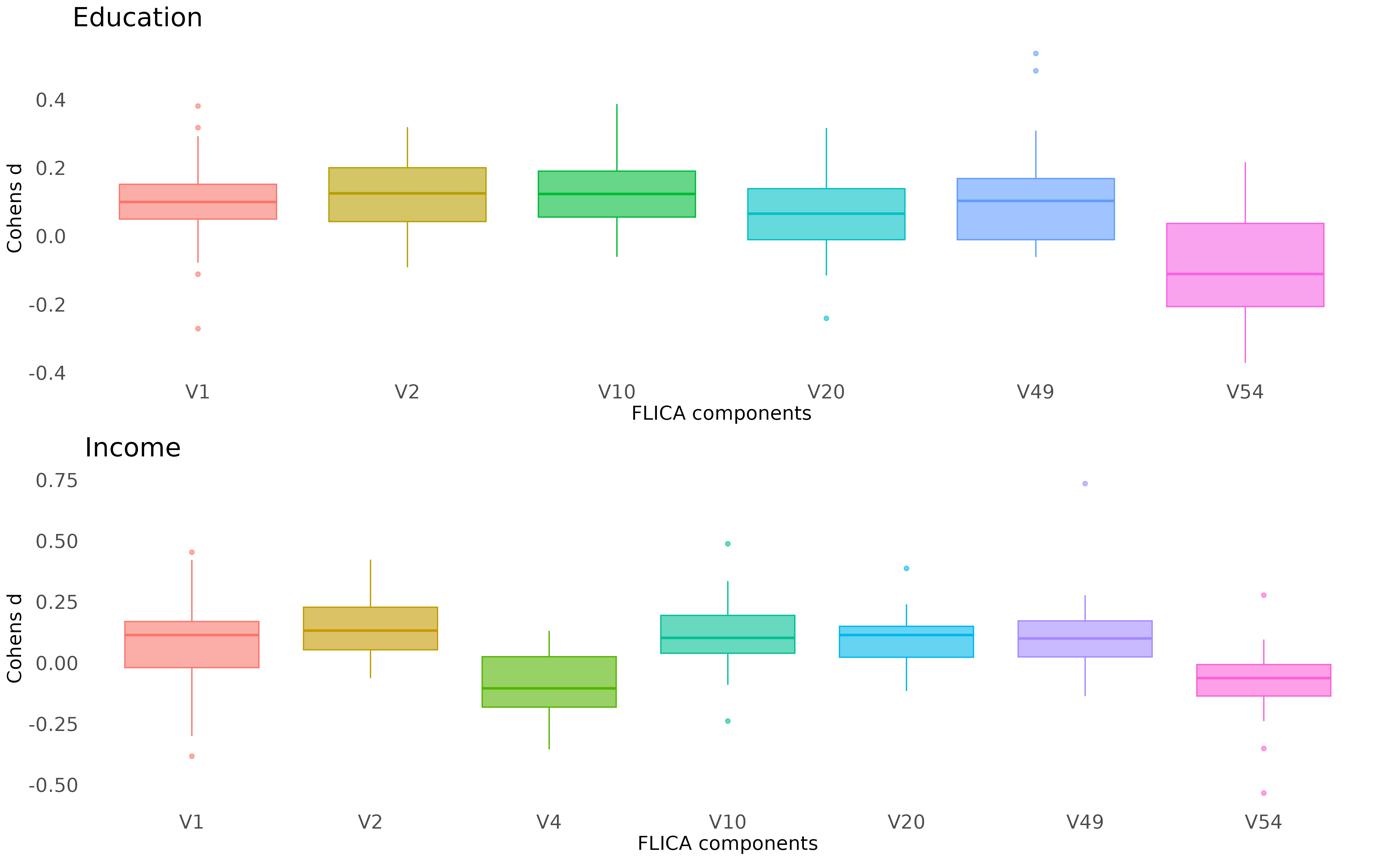

SI Figure 8. The spread of Cohens’ d for components significantly associated with socioeconomic status across scanners. The figure shows box plots of the spread of Cohens’ d for components significantly associated with education or occupation across 27 scanners. Of note, the analysis excluded the random effect of zygosity due to limited variance in this factor within each subset, and two scanners were omitted due to model complexity constraints for these subsets.

*Moderation effects of cortical structure on the parental SES - child p-factor relationship*

Statistical analyses revealed no significant moderation effect of cortical structure on the negative relationship between parental education and the p-factor as presented in SI Table 3. IC54 showed a statistically significant negative moderation effect of the parental income - child p-factor relationship, as presented in SI Table 3.

|  | PARENTAL EDUCATION | | | | | PARENTAL INCOME | | | |
| --- | --- | --- | --- | --- | --- | --- | --- | --- | --- |
| IC | Cohens D | T statistic | | p-value (uncorrected) | p-value (corrected) | Cohens D | T statistic | p-value (uncorrected) | p-value (corrected) |
| 1 | 0.01 | | 0.47 | 0.641 | 0.946 | 0.01 | 0.55 | 0.584 | 0.926 |
| 2 | 0 | | 0.07 | 0.946 | 0.946 | 0.02 | 0.78 | 0.434 | 0.926 |
| 4 | - | | - | - | - | -0.01 | -0.36 | 0.719 | 0.926 |
| 10 | 0.03 | | 1.55 | 0.122 | .683 | 0.04 | 1.82 | 0.068 | 0.479 |
| 20 | -0.01 | | -0.3 | 0.761 | 0.946 | 0 | 0.09 | 0.926 | 0.926 |
| 33 | - | | - | - | - | -0.02 | -1.02 | 0.309 | 0.926 |
| 49 | 0 | | 0.15 | 0.88 | 0.946 | 0.03 | 1.21 | 0.226 | 0.926 |
| 54 | -0.03 | | -1.49 | 0.137 | 0.683 | -0.07 | -3.06 | 0.002* | 0.018* |

SI Table 3. Moderation effects of relevant components on the relationship between parental socioeconomic status (SES) and the p-factor. The table depicts Cohens D, T statistic, and uncorrected- and FDR-corrected p-values from the statistical analyses of the moderation effects of particular component loadings on the relation between parental education, parental income and the p-factor. Significant p-values are marked with Asterix (*).

*Linear association s between parental SES and unimodal global cortical metrics*

To compare our findings to standard unimodal assessments we ran linear mixed effects models testing the association between parental SES and unimodal metrics, separately. Results are presented in SI Table 4.

In general accordance with our multimodal assessments, we found significant positive associations between parental education and youth total surface area and mean GWC. The strongest association was found for surface area. There were no significant associations between parental education and youth cortical thickness or curvature. Similarly, we found significant relations between parental income and youth total surface area and mean GWC, but no significant associations with cortical thickness or curvature.

|  | PARENTAL EDUCATION | | | | | PARENTAL INCOME | | | |
| --- | --- | --- | --- | --- | --- | --- | --- | --- | --- |
| Metric | Cohens D | T statistic | | p-value (uncorrected) | p-value (corrected) | Cohens D | T statistic | p-value (uncorrected) | p-value (corrected) |
| Mean CT | 0.04 | | 1.93 | 0.053 | 0.086 | 0.04 | 1.85 | 0.064 | 0.128 |
| Total area | 0.14 | | 6.19 | >0.001* | >0.001* | 0.13 | 5.87 | >0.001* | >0.001* |
| Mean Curvature | -0.04 | | -1.72 | 0.086 | 0.086 | 0 | -0.03 | 0.973 | 0.973 |
| Mean GWC | 0.08 | | 3.6 | >0.001* | 0.001* | 0.12 | 5.15 | >0.001* | >0.001* |

SI Table 4. Associations between parental socioeconomic status (SES) and unimodal cortical structure. The table depicts Cohens D, T statistic, and uncorrected- and FDR-corrected p-values from the statistical analyses of the associations between parental education, parental income, and mean cortical thickness (CT), total surface area (Total area), mean curvature, and mean grey/white matter contrast (GWC). Significant p-values are marked with Astrix (*).

**Discussion**

The multimodal decomposition of vertex-wise cortical metrics revealed that most of the variance within our data could be explained by global surface area (possibly in part reflecting sex-related differences), closely followed by global GWC. As expected in a sample with a narrow age range, cortical thickness was less dominant, and curvature, which we did not have clear expectations for, hardly fused with the other metrics. FLICA can model unimodality when present, and commonly each cortical metric will show a unimodal global signal pointing to the metrics being partly distinct and correlating within-, before fusing across. It was surprising that cortical thickness did not show such a pattern, possibly pointing to mean thickness being central for development but not for other aspects of youth heterogeneity. In line with our hypothesis multimodal patterns generally captured similar regions across metrics, apart from thickness and curvature not fusing well. Of note, the first component did model higher curvature with thinner cortex (Demirci & Holland, 2022), although with overall weak weighting for both metrics. One possibility for curvature explaining little of the variance within the FLICA decomposition, is that curvature could be the least noisy, thereby being underfitted by FLICA. This does not fully explain our findings as surface area, which dominated our decomposition, showed highly similar noise estimates to thickness which did not. In sum, beyond cortical development, youth studies of demography or behavior, should include multivariate assessments of cortical surface area and GWC, and future studies should clarify such relations to cortical curvature.

**References**

Auchter, A. M., Hernandez Mejia, M., Heyser, C. J., Shilling, P. D., Jernigan, T. L., Brown, S. A., Tapert, S. F., & Dowling, G. J. (2018). A description of the ABCD organizational structure and communication framework. *Developmental Cognitive Neuroscience*, *32*, 8–15. https://doi.org/10.1016/j.dcn.2018.04.003

Demirci, N., & Holland, M. A. (2022). Cortical thickness systematically varies with curvature and depth in healthy human brains. *Human Brain Mapping*, *43*(6), 2064–2084. https://doi.org/10.1002/hbm.25776

Fortin, J.-P., Cullen, N., Sheline, Y. I., Taylor, W. D., Aselcioglu, I., Cook, P. A., Adams, P., Cooper, C., Fava, M., McGrath, P. J., McInnis, M., Phillips, M. L., Trivedi, M. H., Weissman, M. M., & Shinohara, R. T. (2018). Harmonization of cortical thickness measurements across scanners and sites. *NeuroImage*, *167*, 104–120. https://doi.org/10.1016/j.neuroimage.2017.11.024

Karcher, N. R., Barch, D. M., Avenevoli, S., Savill, M., Huber, R. S., Simon, T. J., Leckliter, I. N., Sher, K. J., & Loewy, R. L. (2018). Assessment of the Prodromal Questionnaire–Brief Child Version for Measurement of Self-reported Psychoticlike Experiences in Childhood. *JAMA Psychiatry*, *75*(8), 853. https://doi.org/10.1001/jamapsychiatry.2018.1334
